# Adverse Graft Remodeling Reflects Dynamic Allograft Stress and Predicts Adverse Outcomes After Heart Transplantation

**DOI:** 10.64898/2026.08.25.26361222

**Authors:** Khush Patel, Timothy Pan, Sadeer Al-Kindi, Todd N. Eagar, Guillermo Torre-Amione, Ashrith Guha, Rajul Ranka, Ruli Gao, Arvind Bhimaraj

## Abstract

**BACKGROUND:** Increased left ventricular mass (LVM) at a single time point after heart transplantation (HT) predicts future adverse outcomes. However, dynamic changes in LVM could have better biological relevance and reflect adverse graft remodeling (AGR). The prognostic significance of such serial changes has not been studied.

**METHODS:** Using an automated, electronic health record-based institutional data infrastructure, we studied 439 HT recipients with 5,563 LVM measurements. Separate Bayesian joint models estimated the simultaneous associations of current LVM and its instantaneous rate of change with graft dysfunction (GD) and mortality. A joint-model-derived remodeling score combining patient-specific deviations in LVM and slope was dichotomized to define AGR and non-AGR groups. A mixed-effects analysis of all clinical variables was performed to assess associations with LVM both between and within patients. An independent cohort of 35 patients with 79 surveillance-biopsy RNA-sequencing samples was used to examine early stress-responsive pathways associated with the remodeling score.

**RESULTS:** LVM declined by approximately 7 g/year after transplantation, with regression attenuating over time. Sixty patients (13.7%) had GD, and 75 (17.1%) died. Higher LVM was associated with subsequent GD (hazard ratio [HR] per 10 g, 1.14; 95% credible interval [CrI], 1.02-1.28) and mortality (HR, 1.10; 95% CrI, 1.02-1.19). A more positive LVM slope was associated with GD (HR per 1 g/year, 1.21; 95% CrI, 1.06-1.42) and with cardiac allograft vasculopathy (CAV) grade 2 or 3 (HR, 1.39; 95% Crl, 1.02-1.96). LVM regressed more slowly in the AGR group (-5.8 vs -8.4 g/year), with higher GD (21.0% vs 6.4%) and mortality (24.2% vs 10.0%). Time-updated GD was associated with subsequent death (HR, 8.12; 95% Confidence Interval [CI], 4.67-14.14). Transcriptomic analysis showed enrichment of interferon-mediated signaling and vascular endothelial activation with higher remodeling scores, whereas lower scores were associated with mitochondrial and metabolic processes, ribosome biogenesis, and pathways related to tissue repair and stress responses.

**CONCLUSIONS:** AGR is an easily accessible imaging biomarker that reflects the changes in the allograft in response to various stressors and predicts future adverse outcomes. Discovery of molecular mechanisms of AGR could lead to novel therapies to protect the allograft from chronic rejection.

## Introduction

Imaging-based increased left ventricular mass (LVM) is used as a marker of hypertrophy and organ-level remodeling reflecting a final common structural response to a wide range of stressors on the myocardium. Across multiple non-transplant populations, increased LVM has been established as an independent predictor of cardiovascular morbidity and mortality.(1–3) In heart transplant (HT) recipients, such remodeling occurs within a distinct and complex physiological milieu as a response to various stressors starting from organ retrieval, preservation, and perioperative reperfusion injury to evolving metabolic, immune, and loading conditions throughout the lifetime of the allograft.(4, 5)

Prior studies have suggested that an increased LVM is prevalent and carries independent prognostic significance. (6, 7) While metabolic derangements, pharmacotherapy, and cardiac load can contribute to remodeling, an independent immune mechanism has also been suggested. (8, 9) Despite these foundational observations, prior investigations have largely focused on LVM at an arbitrary single time point. Longitudinal changes in LVM may more accurately reflect the evolving balance between acute response to injury and cumulative allograft adaptation.

Accordingly, we sought to evaluate the prognostic significance of graft remodeling (GR) quantified by both absolute LVM and longitudinal rate of change (slope) utilizing an automated, deeply phenotyped HT dataset from a single center. (10) By integrating standardized echocardiographic assessment with detailed clinical, hemodynamic, immunologic, and vasculopathy characterization, this study aims to extend prior work by moving beyond static measures towards a longitudinal framework of adverse graft remodeling (AGR) associated with clinical outcomes. Further, in a sub-cohort of patients with available RNA-seq data from biopsies, we explored the molecular pathways associated with AGR to gain mechanistic insights into the biological processes underlying AGR and its relationship with allograft injury and dysfunction.

## METHODS

### Study population and data source

We analyzed patients from the J.C. Walter Jr. Transplant Center Precision Registry (TCPR) -Heart.(10) In brief, this is a single-center, automated, multi-source, deeply phenotyped clinical registry built for research in HT. All structured data from the electronic health record (EHR) is automatically available, while specific unstructured data from relevant clinical reports are abstracted using natural language processing. Details of the data query methods are published previously.(10) Echocardiography reports were available from June 2016 through March 2026.

Recipients were eligible if they had at least two echo reports with LVM measurements, including at least one within the first post-transplant year. This yielded a cohort transplanted between June 2015 and March 2026. Patients with graft dysfunction (GD) (defined below) within 90 days after transplantation were excluded to focus on GR that predicts longer-term outcomes. Echocardiograms obtained after the outcome of interest or during the preceding 90 days were excluded from the corresponding event-specific longitudinal dataset to reduce the influence of acute pre-event changes. Because clinically significant cardiac allograft vasculopathy (CAV) may develop over a longer period than the primary cohort’s available follow-up, CAV analyses used a broader cohort with available longitudinal echocardiographic and angiographic surveillance data and did not require an echocardiogram within the first post-transplant year. The study was approved under the transplant center data analysis institutional review board protocol Pro00000587. A sub-cohort of patients (N=35) participated in a consented study where, during their protocol biopsies, a biopsy piece was utilized under a research protocol (Pro00035283) to analyze for bulk RNA sequencing.

### Echocardiogram variables

LVM was calculated programmatically from all available reports using relevant variables based on the American Society of Echocardiography formula(11) : LV mass 0.8 1.04 LVIDD IVSD PWD LVIDD 0.6 g. The LVM index was calculated by dividing LV mass by the recipient’s body surface area.

### Cardiac magnetic resonance validation

Among patients undergoing clinically indicated cardiac magnetic resonance (CMR) imaging, CMR-derived LVM was compared with the nearest echocardiographic measurements obtained within 90 days. Agreement was assessed by intraclass correlation and Bland-Altman analysis.

### Outcomes

GD was defined as an ejection fraction < 50% or a relative decline of at least 25% from the preceding echo(10, 12). Mortality was ascertained from institutional records and United Network for Organ Sharing documentation. CAV was graded by a single experienced transplant cardiologist (AB) reviewing coronary angiograms according to International Society for Heart and Lung Transplantation criteria, modified to exclude graft dysfunction from the grade 3 definition(10, 13). The primary CAV endpoint was the first occurrence of angiographic CAV grades 2 or 3.

### Statistical analysis

Continuous variables were summarized as median (interquartile range) and categorical variables as number (percentage). LVM trajectories were estimated using linear mixed-effects models containing linear and quadratic time after transplantation and patient-specific random intercepts and slopes. Associations of LVM trajectories with GD and mortality were evaluated using separate Bayesian joint models; associations are summarized as hazard ratios with 95% credible intervals(14). For readability, we additionally report a two-sided posterior probability (denoted p), calculated as twice the smaller posterior tail probability relative to the null, which approximates a conventional two-sided p value. The longitudinal sub model included time, time squared, and recipient sex, with patient-specific random intercepts and slopes. The survival sub models used a proportional-hazards model. The models were linked through the current fitted LVM and its instantaneous rate of change, which were included simultaneously. Current-value associations were reported per 10-g higher LVM and slope associations per 1-g/year more positive LVM trajectory. Slope associations are continuous and were not dichotomized at zero; because LVM declined on average throughout follow-up, a more positive slope denotes slower regression as well as absolute growth. Expanded models additionally adjusted for recipient age, donor age, and recipient creatinine at transplantation. Analyses were repeated using LVM indexed to body surface area. A separate joint model evaluated CAV grade 2 or 3. A continuous patient-level LV remodeling score was derived from the GD joint model by combining patient-specific deviations in LVM and LVM slope with their corresponding association coefficients. Patients were divided at the cohort median to illustrate the clinical separation associated with the score. Because GD was used to derive the score, comparisons of GD between remodeling groups were considered descriptive and not an independent test of predictive performance. Mortality was not used in score derivation and was evaluated as a separate outcome.

Candidate factors associated with LVM were evaluated using mixed-effects models adjusted for time, time squared, and recipient sex. Time-varying exposures were decomposed into between-patient and within-patient components. Variables representing nonredundant recipient, donor, hemodynamic, rejection, renal, and metabolic domains were subsequently entered into a multivariable mixed-effects model. In the subset of patients undergoing Molecular Microscope Diagnostic System (MMDx)(15) assessment of endomyocardial biopsy specimens, continuous transcript scores were standardized and linked to each echocardiogram using the closest preceding MMDx assessment within 180 days; same-day assessments were excluded.

The associations of GD and CAV with subsequent mortality were assessed using time-updated Cox models with robust standard errors. Two-sided p values below 0.05 were considered statistically significant. Analyses were performed using R version 4.5.2. Additional cohort-construction rules, exposure-matching procedures, model specifications, sensitivity analyses, and quality-control procedures are provided in the Supplementary Methods.

### Transcriptomic validation cohort

We studied an independent, non-overlapping cohort of 35 heart transplant recipients who had serial echocardiograms and RNA sequencing of surveillance heart biopsies. The analysis included 79 biopsy samples collected 2 to 30 weeks after transplantation, with a median collection time of approximately 10 weeks. We applied the original LV mass model to each patient’s echocardiograms to derive patient-level remodeling scores. Gene-expression associations with the continuous score were tested using limma-voom with empirical Bayes moderation(16, 17), adjusting for biopsy timing and donor sex; duplicate correlation accounted for repeated biopsies within patients. GSEA used MSigDB Hallmark and Gene Ontology collections(18–20), with a false discovery rate <0.05 defining significance. Additional details on cohort selection, sample processing, library preparation, sequencing, read alignment, normalization, and differential expression are available in Supplementary Methods.

## RESULTS

### Study cohort

Of 638 heart transplant recipients during study period, 439 met the prespecified eligibility criteria. These patients contributed 5,563 LVM measurements, with a median of 12 measurements per patient (IQR, 8-16; range, 2-45). The first eligible echocardiogram was performed at a median of 3 days after transplantation (IQR, 2-7), and median LVM across all measurements was 149.5 g (IQR, 123.3-180.0). Baseline recipient, donor, and transplant characteristics are presented in Table 1. During a median follow-up of 3.0 years, GD occurred in 60 patients (13.7%). During a median follow-up of 4.4 years, 75 patients (17.1%) died (Supplementary Figure 1).

**Table 1.** Characteristics of patients with No Adverse Graft Remodeling (non-AGR) and AGR Joint Model-derived LVM.

| Group | Characteristic | Overall<br>N = 439<sup>1</sup> | Non-AGR<br>N = 220<sup>1</sup> | AGR<br>N = 219<sup>1</sup> | p-value<sup>2</sup> |
| --- | --- | --- | --- | --- | --- |
|  | Recipient Characteristics |  |  |  |  |
| Recipient characteristics | Recipient age at transplant, years | 59.0 (52.0, 65.0) | 60.0 (52.0, 66.0) | 58.0 (51.0, 64.0) | 0.093 |
|  | Recipient sex |  |  |  | 0.830 |
|  | Female | 118 (26.9%) | 58 (26.4%) | 60 (27.4%) |  |
|  | Male | 321 (73.1%) | 162 (73.6%) | 159 (72.6%) |  |
|  | Recipient race/ethnicity |  |  |  | 0.185 |
|  | White, Non-Hispanic | 211 (48.1%) | 108 (49.1%) | 103 (47.0%) |  |
|  | Black, Non-Hispanic | 136 (31.0%) | 65 (29.5%) | 71 (32.4%) |  |
|  | Hispanic/Latino | 79 (18.0%) | 37 (16.8%) | 42 (19.2%) |  |
|  | Asian, Non-Hispanic | 10 (2.3%) | 8 (3.6%) | 2 (0.9%) |  |
|  | Amer Ind/Alaska Native, Non-Hispanic | 1 (0.2%) | 0 (0.0%) | 1 (0.5%) |  |
|  | Unknown | 2 (0.5%) | 2 (0.9%) | 0 (0.0%) |  |
|  | Primary diagnosis group |  |  |  | 0.582 |
|  | Congenital heart disease | 20 (4.6%) | 13 (5.9%) | 7 (3.2%) |  |
|  | Dilated cardiomyopathy | 199 (45.4%) | 96 (43.6%) | 103 (47.2%) |  |
|  | Hypertrophic cardiomyopathy | 4 (0.9%) | 1 (0.5%) | 3 (1.4%) |  |
|  | Ischemic cardiomyopathy | 141 (32.2%) | 71 (32.3%) | 70 (32.1%) |  |
|  | Restrictive/infiltrative cardiomyopathy | 54 (12.3%) | 27 (12.3%) | 27 (12.4%) |  |
|  | Valvular heart disease | 2 (0.5%) | 2 (0.9%) | 0 (0.0%) |  |
|  | Other | 18 (4.1%) | 10 (4.5%) | 8 (3.7%) |  |
|  | Recipient blood type |  |  |  | 0.407 |
|  | A | 161 (36.7%) | 85 (38.6%) | 76 (34.7%) |  |
|  | AB | 21 (4.8%) | 10 (4.5%) | 11 (5.0%) |  |
|  | B | 50 (11.4%) | 29 (13.2%) | 21 (9.6%) |  |
|  | O | 207 (47.2%) | 96 (43.6%) | 111 (50.7%) |  |
|  | Recipient BMI at transplant, kg/m<sup>2</sup> | 26.4 (23.5, 29.4) | 25.8 (23.1, 29.0) | 26.9 (23.9, 30.0) | **0.024** |
|  | Urgency status at listing |  |  |  | **0.036** |
|  | Adult Status 1 | 4 (0.9%) | 1 (0.5%) | 3 (1.4%) |  |
|  | Adult Status 2 | 141<br>(32.2%) | 83 (37.9%) | 58 (26.5%) |  |
|  | Adult Status 3 | 12 (2.7%) | 8 (3.7%) | 4 (1.8%) |  |
|  | Adult Status 4 | 82 (18.7%) | 38 (17.4%) | 44 (20.1%) |  |
|  | Adult Status 5 | 13 (3.0%) | 5 (2.3%) | 8 (3.7%) |  |
|  | Adult Status 6 | 43 (9.8%) | 26 (11.9%) | 17 (7.8%) |  |
|  | Status 1A | 51 (11.6%) | 20 (9.1%) | 31 (14.2%) |  |
|  | Status 1B | 55 (12.6%) | 20 (9.1%) | 35 (16.0%) |  |
|  | Status 2 | 37 (8.4%) | 18 (8.2%) | 19 (8.7%) |  |
|  | Missing | 1 | 1 | 0 |  |
|  | Waitlist time, days | 51.0 (21.0,<br>154.0) | 49.0 (18.5,<br>146.5) | 55.0 (23.0,<br>157.0) | 0.184 |
|  | HLA mismatch level |  |  |  | 0.349 |
|  | 1 | 4 (1.0%) | 3 (1.4%) | 1 (0.5%) |  |
|  | 2 | 8 (1.9%) | 4 (1.9%) | 4 (1.9%) |  |
|  | 3 | 43 (10.3%) | 19 (9.0%) | 24 (11.5%) |  |
|  | 4 | 97 (23.2%) | 51 (24.2%) | 46 (22.1%) |  |
|  | 5 | 159<br>(37.9%) | 72 (34.1%) | 87 (41.8%) |  |
|  | 6 | 108<br>(25.8%) | 62 (29.4%) | 46 (22.1%) |  |
|  | Recipient diabetes at listing | 151<br>(34.4%) | 77 (35.0%) | 74 (33.8%) | 0.841 |
|  | Recipient smoking history | 180<br>(41.0%) | 78 (35.5%) | 102 (46.6%) | **0.020** |
|  | Recipient medical condition at transplant |  |  |  | 0.790 |
|  | In Intensive Care Unit | 372<br>(84.9%) | 188 (85.5%) | 184 (84.4%) |  |
|  | Not Hospitalized | 66 (15.1%) | 32 (14.5%) | 34 (15.6%) |  |
|  | Creatinine at time of transplant, mg/dL | 1.3 (1.0, 1.7) | 1.3 (1.0, 1.7) | 1.3 (1.1, 1.7) | 0.324 |
| **Transplant and early post-transplant characteristics** |  |  |  |  |  |
|  | Acute rejection between transplant and discharge |  |  |  | 0.921 |
|  | No | 275<br>(62.8%) | 139 (63.2%) | 136 (62.4%) |  |
|  | Yes, at least one episode treated with anti-rejection agent | 163<br>(37.2%) | 81 (36.8%) | 82 (37.6%) |  |
|  | Length of stay after transplant, days | 21.0 (16.0, 30.0) | 20.0 (16.0, 29.0) | 22.0 (17.0, 30.0) | 0.083 |
|  | Previous other solid-organ transplant | 5 (1.1%) | 3 (1.4%) | 2 (0.9%) | >0.999 |
|  | Multi-organ transplant | 145 | 71 (32.3%) | 74 (33.8%) | 0.761 |
|  |  | (33.0%) |  |  |  |
|  | Simultaneous kidney transplant | 86 | 44 | 42 |  |
|  | Simultaneous liver transplant | 39 | 15 | 24 |  |
|  | Simultaneous lung transplant | 23 | 12 | 11 |  |
|  | Total ischemic time, hours | 3.7 (2.9, 4.1) | 3.7 (2.9, 4.1) | 3.6 (2.9, 4.2) | 0.797 |
|  | **Recipient serologies** |  |  |  |  |
| **Recipient serologies** | Recipient CMV serostatus |  |  |  | >0.999 |
|  | Negative | 151 (34.5%) | 76 (34.5%) | 75 (34.4%) |  |
|  | Positive | 287 (65.5%) | 144 (65.5%) | 143 (65.6%) |  |
|  | Recipient EBV serostatus |  |  |  | 0.098 |
|  | Negative | 55 (12.6%) | 23 (10.5%) | 32 (14.7%) |  |
|  | Positive | 307 (70.1%) | 162 (73.6%) | 145 (66.5%) |  |
|  | Not Done | 70 (16.0%) | 30 (13.6%) | 40 (18.3%) |  |
|  | Unknown | 6 (1.4%) | 5 (2.3%) | 1 (0.5%) |  |
|  | Recipient HBV surface antibody |  |  |  | **0.008** |
|  | Negative | 338 (77.5%) | 159 (72.6%) | 179 (82.5%) |  |
|  | Positive | 91 (20.9%) | 58 (26.5%) | 33 (15.2%) |  |
|  | Not Done | 7 (1.6%) | 2 (0.9%) | 5 (2.3%) |  |
|  | Recipient HBV core antibody |  |  |  | **0.049** |
|  | Negative | 402 (91.8%) | 198 (90.0%) | 204 (93.6%) |  |
|  | Positive | 23 (5.3%) | 17 (7.7%) | 6 (2.8%) |  |
|  | Not Done | 13 (3.0%) | 5 (2.3%) | 8 (3.7%) |  |
|  | Recipient HCV serostatus |  |  |  | 0.253 |
|  | Negative | 424 (96.8%) | 213 (96.8%) | 211 (96.8%) |  |
|  | Positive | 10 (2.3%) | 6 (2.7%) | 4 (1.8%) |  |
|  | Not Done | 3 (0.7%) | 0 (0.0%) | 3 (1.4%) |  |
|  | Unknown | 1 (0.2%) | 1 (0.5%) | 0 (0.0%) |  |
|  | Recipient HIV serostatus |  |  |  | 0.499 |
|  | Negative | 435 (99.3%) | 217 (98.6%) | 218 (100.0%) |  |
|  | Positive | 1 (0.2%) | 1 (0.5%) | 0 (0.0%) |  |
|  | Not Done | 2 (0.5%) | 2 (0.9%) | 0 (0.0%) |  |
|  | **Donor characteristics** |  |  |  |  |
|  | Donor age, years | 29.0 (22.0, 37.0) | 29.0 (22.0, 35.0) | 29.0 (24.0, 39.0) | 0.072 |
|  | Donor sex |  |  |  | **0.017** |
|  | Female | 132<br>(30.1%) | 78 (35.5%) | 54 (24.7%) |  |
|  | Male | 307<br>(69.9%) | 142 (64.5%) | 165 (75.3%) |  |
|  | Donor race/ethnicity |  |  |  | 0.037 |
|  | White, Non-Hispanic | 215<br>(49.0%) | 93 (42.3%) | 122 (55.7%) |  |
|  | Hispanic/Latino | 124<br>(28.2%) | 65 (29.5%) | 59 (26.9%) |  |
|  | Black, Non-Hispanic | 90 (20.5%) | 55 (25.0%) | 35 (16.0%) |  |
|  | Asian, Non-Hispanic | 5 (1.1%) | 3 (1.4%) | 2 (0.9%) |  |
|  | Amer Ind/Alaska Native,<br>Non-Hispanic | 3 (0.7%) | 2 (0.9%) | 1 (0.5%) |  |
|  | Multiracial, Non-Hispanic | 1 (0.2%) | 1 (0.5%) | 0 (0.0%) |  |
|  | Unknown | 1 (0.2%) | 1 (0.5%) | 0 (0.0%) |  |
|  | Donor blood type |  |  |  | 0.496 |
|  | A | 153<br>(34.9%) | 82 (37.3%) | 71 (32.4%) |  |
|  | AB | 7 (1.6%) | 4 (1.8%) | 3 (1.4%) |  |
|  | B | 30 (6.8%) | 17 (7.7%) | 13 (5.9%) |  |
|  | O | 249<br>(56.7%) | 117 (53.2%) | 132 (60.3%) |  |
|  | Donor cause of death |  |  |  | 0.377 |
|  | Anoxia | 133<br>(30.3%) | 75 (34.1%) | 58 (26.5%) |  |
|  | Cerebrovascular/Stroke | 63 (14.4%) | 30 (13.6%) | 33 (15.1%) |  |
|  | Head Trauma | 233<br>(53.1%) | 110 (50.0%) | 123 (56.2%) |  |
|  | Other | 10 (2.3%) | 5 (2.3%) | 5 (2.3%) |  |
|  | Donor history of diabetes | 15 (3.4%) | 7 (3.2%) | 8 (3.7%) | 0.800 |
|  | Donor history of hypertension | 61 (13.9%) | 30 (13.6%) | 31 (14.2%) | 0.891 |
|  | Donor smoking history greater<br>than 20 pack-years | 39 (8.9%) | 20 (9.1%) | 19 (8.7%) | >0.999 |
|  | Donor terminal total bilirubin,<br>mg/dL | 0.7 (0.5,<br>1.3) | 0.7 (0.5, 1.3) | 0.8 (0.5, 1.3) | 0.744 |
|  | Donor terminal blood urea<br>nitrogen, mg/dL | 20.0 (13.0,<br>29.0) | 21.0 (15.0,<br>29.5) | 18.0 (13.0,<br>28.0) | **0.018** |
|  | Donor terminal creatinine,<br>mg/dL | 0.9 (0.7,<br>1.3) | 0.9 (0.7, 1.3) | 0.9 (0.7, 1.2) | 0.742 |
|  | Donor clinical infection | 332<br>(75.6%) | 168 (76.4%) | 164 (74.9%) | 0.740 |
|  | Donor weight, kg | 82.0 (70.2,<br>99.8) | 81.1 (69.4,<br>97.8) | 83.0 (71.2,<br>100.0) | 0.271 |
|  | Donor height, cm | 175.0<br>(168.0,<br>180.0) | 173.0 (167.3,<br>180.0) | 175.0 (168.0,<br>180.0) | 0.088 |
| **Donor serologies** | Donor CMV serostatus |  |  |  | 0.784 |
|  | Negative | 119<br>(27.1%) | 58 (26.4%) | 61 (27.9%) |  |
|  | Positive | 317<br>(72.2%) | 161 (73.2%) | 156 (71.2%) |  |
|  | Not Done | 3 (0.7%) | 1 (0.5%) | 2 (0.9%) |  |
|  | Donor HBV core antibody |  |  |  | 0.503 |
|  | Negative | 430<br>(97.9%) | 214 (97.3%) | 216 (98.6%) |  |
|  | Positive | 9 (2.1%) | 6 (2.7%) | 3 (1.4%) |  |
|  | Donor HCV antibody |  |  |  | 0.338 |
|  | Negative | 430<br>(97.9%) | 217 (98.6%) | 213 (97.3%) |  |
|  | Positive | 9 (2.1%) | 3 (1.4%) | 6 (2.7%) |  |
|  | Donor HIV serostatus |  |  |  | >0.999 |
|  | Negative | 387<br>(88.2%) | 194 (88.2%) | 193 (88.1%) |  |
|  | Not Done | 52 (11.8%) | 26 (11.8%) | 26 (11.9%) |  |
|  | Echocardiographic surveillance |  |  |  |  |
|  | LV mass measurements per patient |  | 12 (8–15) | 13 (8–17) | 0.33 |
|  | Measurements in first year |  | 8 (6–10) | 8 (6–11) | 0.62 |
|  | First measurement, days |  | 3 (2–5) | 4 (2–7) | 0.067 |
|  | Last measurement, years |  | 2.39<br>(1.04–5.17) | 2.40<br>(0.86–5.03) | 0.46 |
|  | Observation span, years |  | 2.32<br>(1.02–5.14) | 2.38<br>(0.76–5.00) | 0.38 |
|  | Observed graft-dysfunction follow-up, days | 1,105.0<br>(457.0, 2,103.0) | 1,140.0<br>(498.5, 2,203.5) | 1,095.0<br>(447.0, 2,051.0) | 0.405 |
|  | Observed survival follow-up, days | 1,620.0<br>(803.0, 2,610.0) | 1,496.5<br>(796.0, 2,584.5) | 1,671.0<br>(808.0, 2,627.0) | 0.705 |
| <sup>1</sup>Median (Q1, Q3); n (%) |  |  |  |  |  |
| <sup>2</sup>Wilcoxon rank sum test; Fisher's exact test; Fisher's Exact Test for Count Data with simulated p-value (based on 1e+05 replicates) |  |  |  |  |  |

### Natural history of LVM change

LVM declined initially after transplantation, with the rate of decline attenuating over time. In the mixed-effects model, estimated mean LVM at transplantation was 133.3 g and initially declined by 7.1 g/year, with the rate of decline slowing over time (quadratic term, +0.66 g/year²; both p<0.001). In the recipient-sex model, male recipients had an average LV mass 34.7 g higher than female recipients throughout follow-up (p<0.001).

In a separate model categorizing donor-recipient sex pairing into four groups, adjusted LVM was highest in male donor-male recipient pairs and lowest in female donor-female recipient pairs. Among male recipients, receipt of a female-donor heart was associated with 25.23 g lower LVM than receipt of a male-donor heart (p<0.001). Among female recipients, receipt of a male-donor heart was associated with 14.11 g higher LVM than receipt of a female-donor heart (p=0.009). There was no evidence that LVM trajectories differed over time by donor-recipient sex pairing (likelihood-ratio p=0.357; Figure 1).

**Figure 1.**
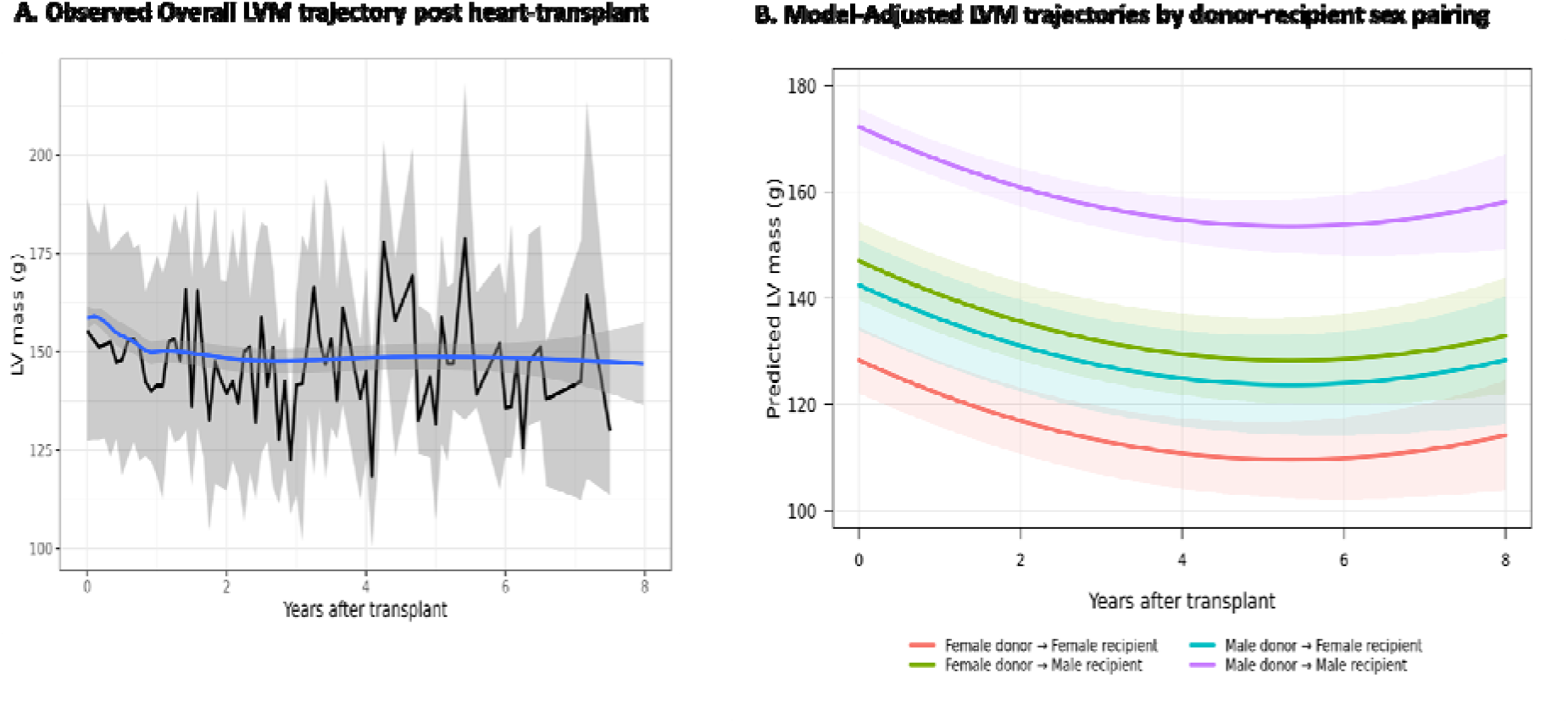
Post-transplant left ventricular mass trajectories overall and according to donor-recipient sex pairing. **A**, Descriptive overall LV-mass trajectory in the primary graft-dysfunction cohort. Measurements were grouped into approximately monthly intervals, and intervals containing fewer than 10 patients were excluded. The black line represents the median LV mass within each interval, and the gray ribbon represents the corresponding interquartile range. The blue line represents a locally estimated scatterplot smoothing curve fitted to all individual LV-mass observations using a span of 0.6; the blue shaded band represents its 95% confidence interval. **B**, Population-average predicted LV-mass trajectories according to donor-recipient sex pairing, estimated from a quadratic linear mixed-effects model containing linear and quadratic time after transplantation and four-category donor-recipient sex pairing, with patient-specific random intercepts and random slopes for linear time. Shaded bands represent 95% confidence intervals for the estimated mean trajectories. Interactions of sex pairing with linear and quadratic time did not improve model fit by likelihood-ratio testing (*p*=0.357), indicating no evidence that the trajectories differed over time among pairing groups.

### LVM trajectory and clinical outcomes

Separate recipient-sex-adjusted joint models assessed GD and death (Figure 2). Higher current LVM was associated with subsequent GD (HR per 10 g, 1.14; 95% CrI, 1.02-1.28) and mortality (HR per 10 g, 1.10; 95% CrI, 1.02-1.19). A more positive LVM slope, indicating slower regression, absence of regression, or progression, was independently associated with GD (HR per 1 g/year, 1.21; 95% CrI, 1.06-1.42), but not mortality (HR per 1 g/year, 1.04; 95% CrI, 0.97-1.13). Results were similar after additional adjustment for recipient age, donor age, and recipient creatinine at transplantation. Associations were preserved when LVM was indexed to recipient body surface area. Higher current LVM index was associated with GD (HR per 10 g/m², 1.35; 95% CrI, 1.04-1.79) and mortality (HR, 1.30; 95% CrI, 1.08-1.57), and a more positive indexed slope with GD (HR per 1 g/m²/year, 1.41; 95% CrI, 1.14-1.76) and mortality (HR, 1.16; 95% CrI, 1.01-1.37).

**Figure 2.**
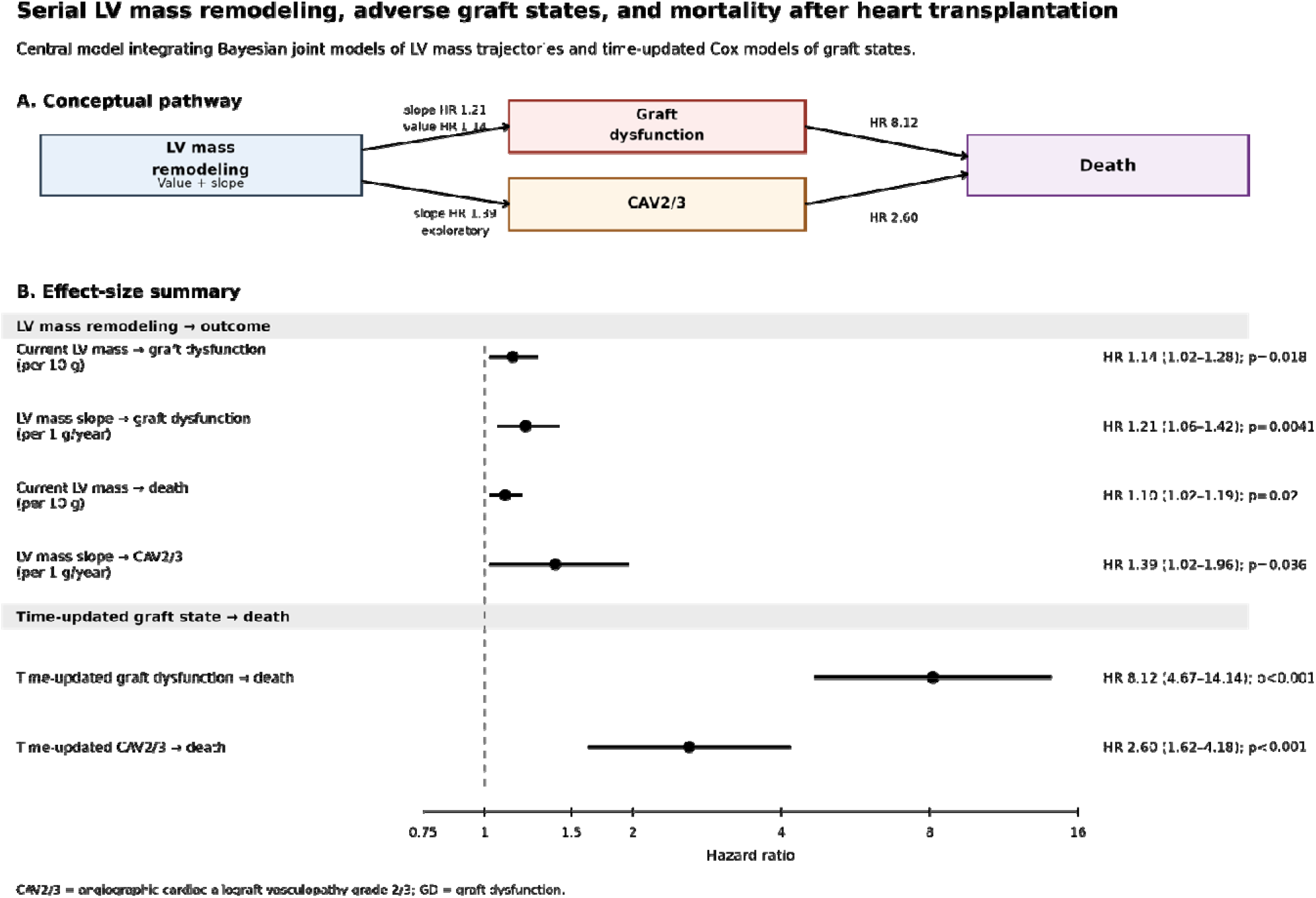
Serial LV mass remodeling, adverse graft states, and mortality after heart transplantation. **A**, Conceptual summary of observed associations. Bayesian joint models linked the underlying longitudinal LV mass trajectory to subsequent outcomes through the current fitted LV mass and its instantaneous rate of change; current value and slope were estimated simultaneously. Higher current LV mass and a more positive LV mass slope were associated with subsequent graft dysfunction, whereas a more positive slope was associated with CAV grade 2 or 3 in an exploratory analysis. Time-updated graft dysfunction and CAV grade 2 or 3 were each associated with subsequent mortality. Arrows summarize statistical associations and should not be interpreted as establishing a causal or obligatory sequence. **B**, Forest plot of effect estimates. Joint-model hazard ratios are reported per 10-g higher current LV mass or per 1-g/year more positive LV mass slope, with 95% credible intervals. Time-updated Cox-model hazard ratios are shown with 95% confidence intervals. The vertical dashed line indicates a hazard ratio of 1. CAV2/3 indicates angiographic cardiac allograft vasculopathy grade 2 or 3; GD, graft dysfunction; HR, hazard ratio

### Adverse graft remodeling (AGR) phenotype

A remodeling score incorporating each patient’s deviation from expected LVM and LVM slope classified 219 patients as having adverse graft remodeling (AGR) and 220 as not AGR (non-AGR group). LVM declined in both groups; however, patients in the AGR group had a higher estimated LVM at transplantation (139.9 versus 126.6 g) with a slower regression compared to the non-AGR group (−5.8 versus −8.4 g/year; Figure 3). Because the remodeling score combined deviations in both LVM level and slope, the two groups differed on both components by construction, and the relative contribution of each was not separately quantified. Notably, however, mean LVM declined in both groups, so adverse remodeling represented attenuated reverse remodeling rather than absolute LVM growth. Baseline characteristics by remodeling group are presented in Table 1. Most baseline characteristics were not significantly different except higher BMI (26.9 vs 25.8; p=0.024), higher prevalence of smoking history (47% vs 35%, p=0.020), higher prevalence of male donors (75% vs 64%, p=0.017) and a lower blood urea nitrogen in donor (18 vs 21 mg/dL, p=0.018) in the AGR group. AGR and non-AGR groups had similar numbers of echocardiograms per patient (median, 13 versus 12; p=0.326), measurements during the first post-transplant year (median, 8 versus 8; p=0.619), observation spans (median, 2.38 versus 2.32 years; p=0.378), and annual imaging frequencies (median, 5.8 versus 5.0 echocardiograms/year; p=0.068). The timing of the first and last eligible echocardiograms also did not differ significantly between groups.

**Figure 3.**
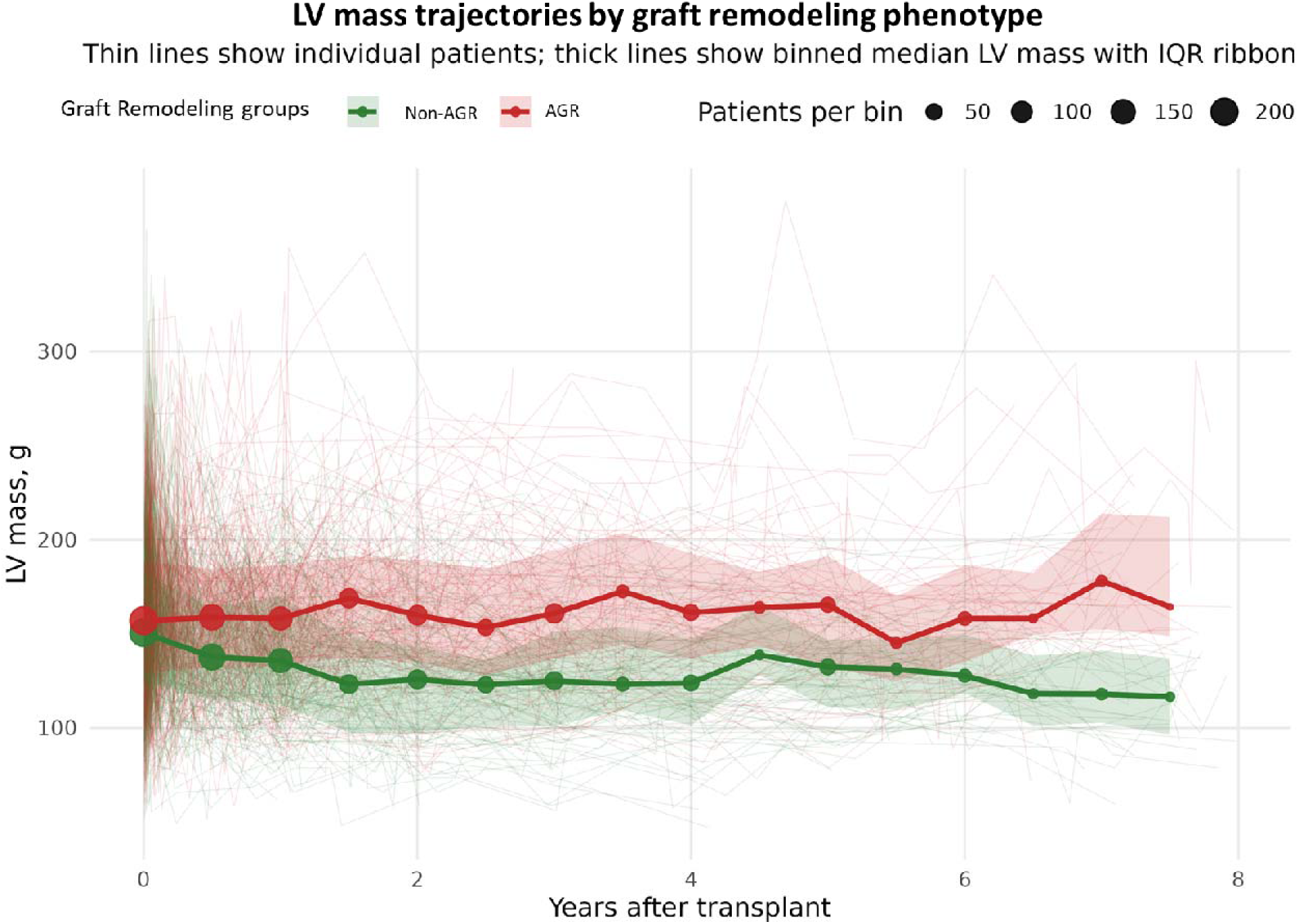
Left ventricular mass trajectories according to graft-remodeling phenotype. Serial LV mass measurements are shown for patients with adverse graft remodeling (AGR) (red; *n*=219) and non-adverse graft remodeling (non-AGR) (green; *n*=220). Thin lines represent individual patient trajectories; thick lines and points represent binned median LV mass, shaded ribbons represent the interquartile range, and point size is proportional to the number of patients contributing to each time bin. LV mass declined in both groups, but the model-estimated rate of regression was slower in the AGR group than in the non-AGR group (−5.8 versus −8.4 g/year). Thus, adverse remodeling primarily reflected impaired reverse remodeling rather than absolute growth in LV mass. The phenotype was defined at the cohort median of a score combining patient-specific deviations in LV mass level and slope, weighted by their associations with graft dysfunction. Group separation is presented to illustrate the clinical magnitude of the modeled associations and does not constitute independent validation.

Because the remodeling score was derived from the GD model, the separation in GD outcomes was expected by construction and is presented descriptively. GD occurred in 46 patients in AGR group (21.0%) compared with 14 patients in the non-AGR group (6.4%; HR, 3.51; 95% CI, 1.93-6.38; p<0.001; Figure 4A). Death occurred in 53 of 219 patients in AGR group (24.2%) compared with 22 of 220 patients in the non-AGR group (10.0%; HR, 2.39; 95% CI, 1.46-3.94; p<0.001; Figure 4B).

**Figure 4.**
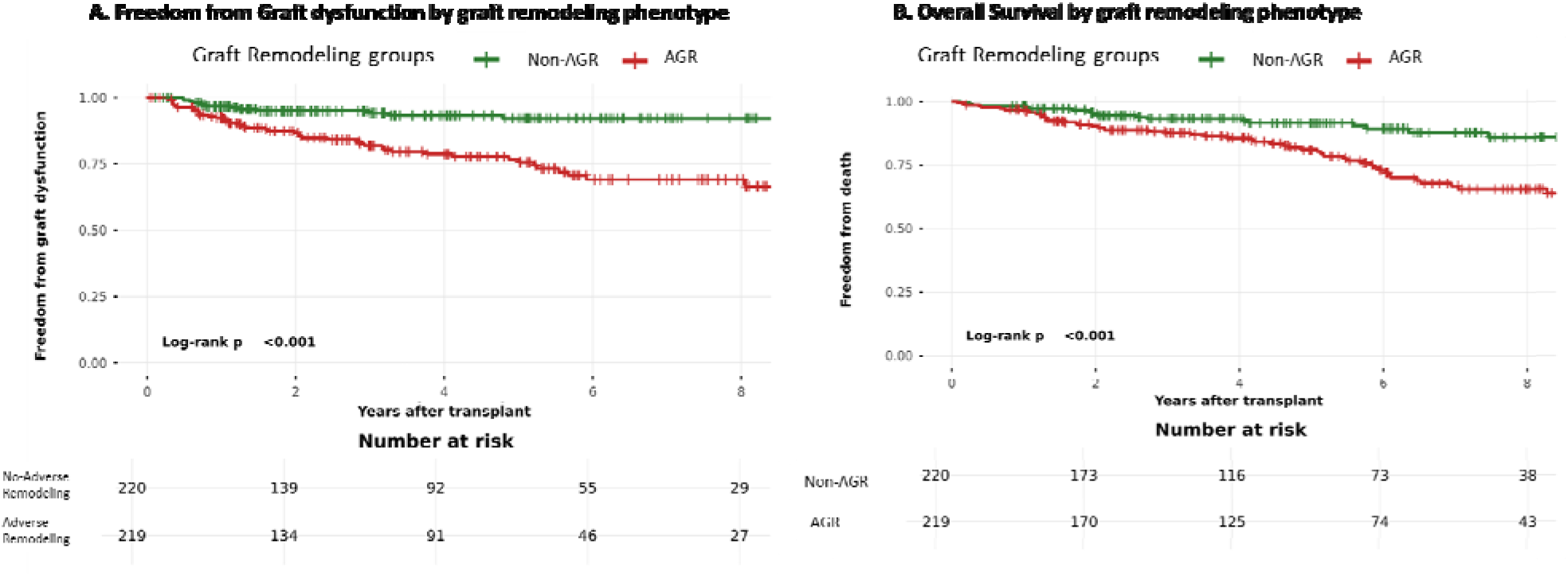
Clinical outcomes according to joint-model-derived graft-remodeling phenotype. Kaplan-Meier curves compare patients with adverse graft remodeling (AGR) (red; n=219) and non-adverse graft remodeling (non-AGR) (green; n=220). Panel A shows freedom from first graft-dysfunction event, and Panel B shows overall survival. Tick marks indicate censored observations, and tables below each panel show the numbers of patients at risk. Patients with AGR had a higher incidence of graft dysfunction than those with non-AGR remodeling (46 of 219 [21.0%] versus 14 of 220 [6.4%]; HR, 3.51; 95% CI, 1.93-6.38; log-rank p<0.001) and higher mortality (53 of 219 [24.2%] versus 22 of 220 [10.0%]; HR, 2.39; 95% CI, 1.46-3.94; log-rank p<0.001). The remodeling phenotype was defined at the cohort median as a score combining patient-specific deviations in LV mass level and slope, weighted by their associations with graft dysfunction. These comparisons illustrate the clinical magnitude of the modeled associations and do not constitute independent prognostic validation.

### Angiographic CAV

The exploratory CAV joint-model analysis included 294 patients with available longitudinal LVM measurements and angiographic CAV grading from 813 coronary angiograms (3,611 eligible LVM measurements and 42 CAV grade 2 or 3 events). Current LVM was not significantly associated with CAV (HR per 10 g, 0.87; 95% CrI, 0.73-1.01; p=0.068), but a more positive LVM slope was associated with subsequent angiographic CAV grade 2 or 3 (HR per 1 g/year, 1.39; 95% CrI, 1.02-1.96; p=0.036; (Figure 2). Our center CAV surveillance protocol uses invasive angiography starting year 5 from HT.

### Time-updated associations of GD and CAV with mortality

When modeled as a time-updated exposure, GD was independently associated with subsequent mortality (HR, 8.12; 95% CI, 4.67-14.14; p<0.001; Adjusted for remodeling score-HR, 7.88; 95% CI, 4.52-13.74; p<0.001). (Figure 2). In the angiographic-surveillance cohort, time-updated CAV grade 2 or 3 was associated with subsequent mortality (HR, 2.60; 95% CI, 1.62-4.18; p<0.001; 308 patients and 94 deaths).

### Adjusted candidate correlates of AGR

A mixed-effect analysis of clinical variables that are associated with LVM between patients showed many established donor and recipient characteristics that are expected to reflect sex and body size-based differences (Table 2 and Figure 5). LVM was higher with male recipients compared to female, greater donor height (+1.21 g per 1 cm) and donor weight (+0.57 g per 1 kg), higher recipient BMI at transplantation (+1.74 g per 1 kg/m²), older donor age (+0.56 g per 1 year), and lower with older recipient age (−0.35 g per 1 year; p=0.008; all other p<0.001). Donor terminal creatinine was associated with a higher LVM (+4.67 g per 1 mg/dL; p<0.001). Every 10-mmHg increase in pulse pressure, systolic blood pressure, and mean arterial pressure after transplant was associated with an 11.16-g increase (95% CI, 7.88-14.44; p<0.001), an 8.34-g increase (95% CI, 5.35-11.33; p<0.001), and a 4.45-g increase (95% CI, 0.22-8.68; p=0.039) in LVM, respectively. Other laboratory parameters that were significant were anion gap, creatinine, protein and albumin/globulin ratio. (Table 2) Of note, the immunosuppression levels and cardiac biomarkers did not differentiate patients with higher LVM. Of the many rejection-based variables, the only measure that met significance was the Endothelial DSA selective transcript of the MMDx test (11.88 g (95% CI: 3.64-20.11) per 1 SD higher score, p=0.005).

**Figure 5.**
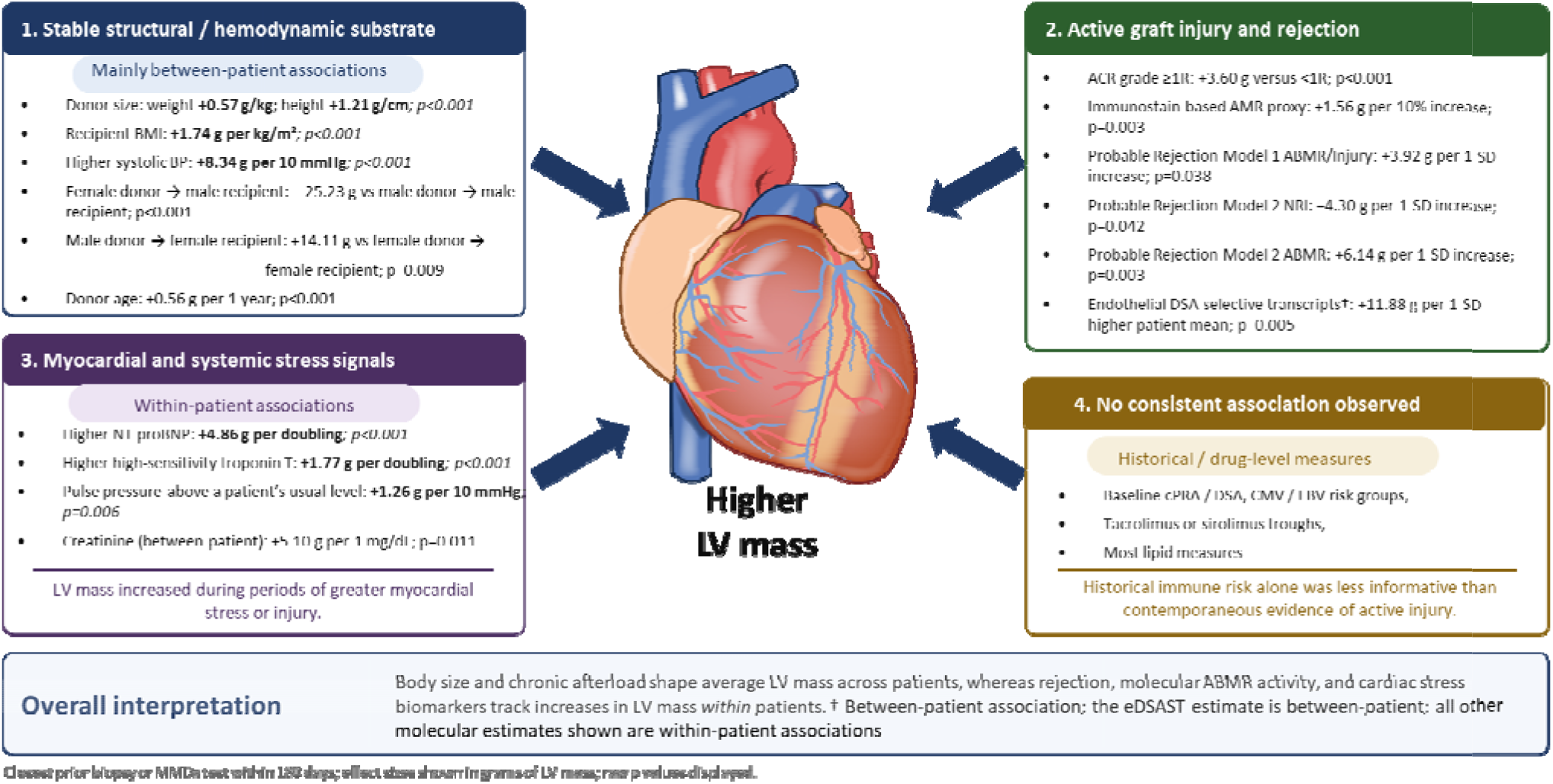
Factors associated with LV mass after heart transplantation. Conceptual summary of the determinants of allograft LV mass, grouped by the process they reflect. Effect sizes are adjusted differences in LV mass (g) from linear mixed-effects models; exposures were matched to the closest preceding biopsy, laboratory or molecular measurement within the specified window. Between-patient effects compare patients with different average exposure; within-patient effects reflect deviation from a patient’s own mean. **Abbreviations:** ABMR, antibody-mediated rejection; ACR, acute cellular rejection; BMI, body mass index; CMV, cytomegalovirus; cPRA, calculated panel-reactive antibody; DSA, donor-specific antibody; EBV, Epstein-Barr virus; eDSAST, endothelial DSA-selective transcripts; LV, left ventricular; MMDx, Molecular Microscope Diagnostic System; NRI, non-rejection injury; NT-proBNP, N-terminal pro-B-type natriuretic peptide; SD, standard deviation.

**Table 2.** Between-patient determinants of left ventricular mass after heart transplantation. Estimates are adjusted differences in LV mass (g) from linear mixed-effects models containing linear and quadratic time after transplantation and recipient sex, except where recipient sex was incorporated into the exposure definition. All comparisons shown are BETWEEN patients: they compare patients whose average level of an exposure was higher with patients whose average level was lower, and answer the question, "Do patients who run higher on this variable have a larger left ventricular mass than other patients?" Donor, recipient, and baseline immunologic characteristics are fixed for a given patient and are therefore between-patient comparisons by definition. The corresponding WITHIN-patient comparisons, in which each patient serves as his or her own control, are reported in Supplementary Tables S1-S4.

| Variable | Scale / contrast | Observations | Patients | Adjusted difference in LV mass, g | 95% CI | p value |
| --- | --- | --- | --- | --- | --- | --- |
| **Donor and recipient structural characteristics** |  |  |  |  |  |  |
| Donor weight | per 1 kg | 5,563 | 439 | **0.57** | **0.45 to 0.70** | **<0.001** |
| Donor height | per 1 cm | 5,563 | 439 | **1.21** | **0.90 to 1.51** | **<0.001** |
| BMI at listing | per 1 kg/m<sup>2</sup> | 5,356 | 420 | **1.98** | **1.40 to 2.56** | **<0.001** |
| BMI at transplant | per 1 kg/m<sup>2</sup> | 5,552 | 437 | **1.74** | **1.14 to 2.34** | **<0.001** |
| Donor terminal creatinine | per 1 mg/dL | 5,563 | 439 | **4.67** | **2.36 to 6.98** | **<0.001** |
| Donor age | per 1 year | 5,563 | 439 | **0.56** | **0.26 to 0.85** | **<0.001** |
| Donor-minus-recipient age difference | per 10 years | 5,563 | 439 | **4.52** | **2.59 to 6.46** | **<0.001** |
| Donor hematocrit | per 1 % | 5,563 | 439 | **0.93** | **0.34 to 1.53** | **0.002** |
| Donor terminal AST | per 1 U/L | 5,563 | 439 | **-0.027** | **-0.047 to -0.007** | **0.008** |
| Recipient age at transplant | per 1 year | 5,563 | 439 | **-0.35** | **-0.60 to -0.09** | **0.008** |
| Donor-recipient sex pairing | Female donor → male recipient vs male donor → male recipient | 5,563 | 439 | **-25.23** | **-33.18 to -17.27** | **<0.001** |
| Donor-recipient sex pairing | Male donor → female recipient vs female donor → female recipient | 5,563 | 439 | **14.11** | **3.60 to 24.61** | **0.009** |
| Donation after circulatory death | DCD vs DBD | 5,563 | 439 | -2.85 | -18.62 to 12.93 | 0.723 |
| Total ischemic time | per 1 hour | 5,552 | 437 | 0.86 | -1.69 to 3.42 | 0.507 |
| Donor cause of death | Cerebrovascular/stroke vs anoxia | 5,563 | 439 | 2.67 | -6.03 to 11.37 | 0.546 |
| Donor cause of death | Head trauma vs anoxia | 5,563 | 439 | 0.27 | -5.92 to 6.45 | 0.933 |
| Donor cause of death | Other vs anoxia | 5,563 | 439 | 11.12 | -7.52 to 29.76 | 0.242 |
| **Hemodynamic measures** |  |  |  |  |  |  |
| Pulse pressure | per 10 mmHg | 4,221 | 425 | **11.16** | **7.88 to 14.44** | **<0.001** |
| Systolic blood pressure | per 10 mmHg | 4,221 | 425 | **8.34** | **5.35 to 11.33** | **<0.001** |
| Mean arterial pressure | per 10 mmHg | 4,221 | 425 | **4.45** | **0.22 to 8.68** | **0.039** |
| **Laboratory, cardiac biomarker, metabolic, lipid, and immunosuppressant measures** |  |  |  |  |  |  |
| Sodium | per 1 mmol/L | 5,523 | 439 | 0.50 | -0.90 to 1.90 | 0.483 |
| Potassium | per 1 mmol/L | 5,530 | 439 | -5.14 | -16.00 to 5.71 | 0.352 |
| Chloride | per 1 mmol/L | 5,523 | 439 | -0.92 | -1.96 to 0.13 | 0.086 |
| CO<sub>2</sub> | per 1 mmol/L | 5,523 | 439 | -0.72 | -2.40 to 0.97 | 0.405 |
| Anion gap | per 1 mmol/L | 5,514 | 439 | **3.70** | **2.12 to 5.28** | **<0.001** |
| Blood urea nitrogen | per 10 mg/dL | 5,523 | 439 | 2.49 | -0.22 to 5.19 | 0.071 |
| Creatinine | per 1 mg/dL | 5,523 | 439 | **5.10** | **1.17 to 9.04** | **0.011** |
| Glucose | per 10 mg/dL | 5,523 | 439 | 0.57 | -0.38 to 1.51 | 0.237 |
| Calcium | per 1 mg/dL | 5,523 | 439 | 4.40 | -2.38 to 11.18 | 0.203 |
| Total protein | per 1 g/dL | 5,495 | 439 | **6.77** | **1.97 to 11.57** | **0.006** |
| Albumin | per 1 g/dL | 5,494 | 439 | 5.40 | -1.94 to 12.74 | 0.149 |
| Albumin/globulin ratio | per 1 unit | 4,649 | 439 | **-17.58** | **-30.55 to -4.62** | **0.008** |
| Alkaline phosphatase | per 10 U/L | 5,494 | 439 | 0.37 | -0.16 to 0.89 | 0.168 |
| AST | per 10 U/L | 5,494 | 439 | -0.23 | -1.14 to 0.67 | 0.612 |
| ALT | per 10 U/L | 5,495 | 439 | -0.19 | -1.03 to 0.64 | 0.646 |
| Total bilirubin | per 1 mg/dL | 5,495 | 439 | -0.45 | -5.25 to 4.36 | 0.855 |
| Tacrolimus trough | per 1 ng/mL | 2,595 | 352 | -0.28 | -1.62 to 1.07 | 0.688 |
| Sirolimus trough | per 1 ng/mL | 98 | 26 | 4.23 | -0.16 to 8.61 | 0.058 |
| NT-proBNP | per doubling | 986 | 238 | 1.94 | -0.12 to 4.01 | 0.065 |
| High-sensitivity troponin T | per doubling | 1,590 | 284 | -0.40 | -2.13 to 1.33 | 0.650 |
| Creatine kinase | per doubling | 668 | 144 | -0.59 | -4.14 to 2.95 | 0.741 |
| Total cholesterol | per 10 mg/dL | 3,437 | 413 | 0.39 | -0.51 to 1.28 | 0.398 |
| Triglycerides | per doubling | 3,565 | 417 | 0.09 | -5.07 to 5.25 | 0.973 |
| HDL cholesterol | per 10 mg/dL | 3,043 | 405 | -0.83 | -2.68 to 1.01 | 0.375 |
| LDL cholesterol | per 10 mg/dL | 3,026 | 405 | 1.04 | -0.16 to 2.23 | 0.089 |
| Hemoglobin A1c | per 1 percentage point | 3,341 | 408 | -0.65 | -3.36 to 2.07 | 0.641 |
| **Rejection and alloimmune measures** |  |  |  |  |  |  |
| Acute cellular rejection | Closest preceding biopsy ACR grade $\geq 1R$ vs $<1R$ | 5,533 | 439 | 3.51 | -8.82 to 15.84 | 0.576 |
| C4d immunostain (AMR proxy) | per 10 percentage points | 5,461 | 439 | -1.16 | -7.59 to 5.27 | 0.724 |
| cPRA class I at transplant | per 10 percentage points | 3,131 | 293 | -0.20 | -1.86 to 1.45 | 0.809 |
| cPRA class II at transplant | per 10 percentage points | 3,131 | 293 | -0.79 | -2.13 to 0.56 | 0.251 |
| DSA class I at transplant | Positive vs negative | 3,131 | 293 | 1.93 | -4.48 to 8.35 | 0.554 |
| DSA class II at transplant | Positive vs negative | 3,131 | 293 | -0.46 | -7.18 to 6.26 | 0.893 |
| EBV high risk | D+/R- vs not D+/R- | 4,319 | 354 | -0.41 | -8.52 to 7.70 | 0.921 |
| CMV high risk | D+/R- vs not D+/R- | 5,522 | 435 | 1.94 | -4.45 to 8.33 | 0.551 |
| **Molecular (MMDx) transcript scores — exploratory subgroup** |  |  |  |  |  |  |
| Probable Rejection Model 1 TCMR/Injury | per 1 SD | 497 | 80 | -2.61 | -14.55 to 9.34 | 0.665 |
| Probable Rejection Model 1 NRI | per 1 SD | 497 | 80 | -3.04 | -14.49 to 8.41 | 0.599 |
| Probable Rejection Model 1 ABMR/Injury | per 1 SD | 497 | 80 | -0.55 | -11.23 to 10.13 | 0.919 |
| Probable Rejection Model 2 TCMR | per 1 SD | 497 | 80 | 8.20 | -7.50 to 23.89 | 0.302 |
| Probable Rejection Model 2 NRI | per 1 SD | 497 | 80 | 2.91 | -10.15 to 15.96 | 0.659 |
| Probable Rejection Model 2 ABMR | per 1 SD | 497 | 80 | 1.06 | -14.92 to 17.04 | 0.895 |
| Enzyme (ADAMDEC1) | per 1 SD | 497 | 80 | -2.25 | -13.42 to 8.91 | 0.689 |
| Checkpoint (CTLA4) | per 1 SD | 497 | 80 | 0.64 | -9.31 to 10.59 | 0.898 |
| Cytokine (CXCL13) | per 1 SD | 497 | 80 | -4.59 | -17.70 to 8.52 | 0.488 |
| DSA-selective (DSAST) | per 1 SD | 497 | 80 | 6.68 | -0.85 to 14.22 | 0.081 |
| IFNG inducible (GRIT) | per 1 SD | 497 | 80 | 2.19 | -5.75 to 10.13 | 0.585 |
| Heart transcripts (HT1) | per 1 SD | 497 | 80 | -4.40 | -12.50 to 3.69 | 0.282 |
| Interferon gamma (IFNG) | per 1 SD | 497 | 80 | -2.76 | -12.10 to 6.57 | 0.557 |
| Injury transcripts (IRRAT) | per 1 SD | 497 | 80 | 5.75 | -2.50 to 14.01 | 0.169 |
| NK cell burden (NKB) | per 1 SD | 497 | 80 | 4.86 | -3.22 to 12.94 | 0.235 |
| PC1 | per 1 SD | 497 | 80 | 2.14 | -6.13 to 10.41 | 0.608 |
| PC2 | per 1 SD | 497 | 80 | 6.64 | -1.91 to 15.19 | 0.126 |
| PC3 | per 1 SD | 497 | 80 | 2.00 | -5.89 to 9.89 | 0.615 |
| Cytotoxic T cell transcripts (QCAT) | per 1 SD | 497 | 80 | 3.77 | -4.31 to 11.85 | 0.356 |
| Macrophage transcripts (QCMAT) | per 1 SD | 497 | 80 | 2.03 | -6.11 to 10.17 | 0.621 |
| Rejection Model 1 NRI | per 1 SD | 497 | 80 | -2.97 | -10.90 to 4.95 | 0.458 |
| Rejection Model 1 TCMR/injury | per 1 SD | 497 | 80 | -1.50 | -11.73 to 8.74 | 0.772 |
| Rejection Model 1 ABMR/injury | per 1 SD | 497 | 80 | 4.20 | -3.32 to 11.72 | 0.270 |
| Rejection Model 2 NRI | per 1 SD | 497 | 80 | -2.80 | -10.81 to 5.20 | 0.487 |
| Rejection Model 2 TCMR | per 1 SD | 497 | 80 | -1.10 | -13.63 to 11.43 | 0.862 |
| Rejection Model 2 ABMR | per 1 SD | 497 | 80 | 4.48 | -3.05 to 12.02 | 0.240 |
| Rejection Model 2 Injury | per 1 SD | 497 | 80 | 0.14 | -8.18 to 8.46 | 0.973 |
| ROBO4 | per 1 SD | 497 | 80 | 6.26 | -1.15 to 13.67 | 0.097 |
| T cell burden (TCB) | per 1 SD | 497 | 80 | 1.61 | -6.35 to 9.58 | 0.688 |
| Endothelial DSA-selective (eDSAST) | per 1 SD | 497 | 80 | **11.88** | **3.64 to 20.11** | **0.005** |

Reflecting a dynamic nature of LVM association in a post HT patients’ lifetime, many more variables were associated with changes in LVM in the within-patient comparison (Supplementary tables S1-S4, Figure 5). While various hemodynamic and laboratory values were significantly associated with LVM within a patient, they contributed modestly. NT-proBNP doubling had the largest positive association with an increase of 4.86g (95% CI, 3.44-6.29; p<0.001) and high-sensitivity Troponin T doubling was associated with a 1.77 g increase (95% CI, 0.90-2.65; p<0.001) (Supplementary table S1). Immune injury was associated with contemporaneous increases in LVM in the within patient comparison (Supplementary table S2). Compared with periods without rejection, LVM was 3.60 g higher when the closest biopsy within the preceding 180 days showed acute cellular rejection grade ≥1R (95% CI, 1.50-5.69; p<0.001). Each 10-percentage-point increase in C4d Immunostaining positivity, used as an antibody-mediated rejection proxy, was associated with 1.56 g higher LVM (95% CI, 0.53-2.60; p=0.003). A subgroup analysis for those patients who had at least 2 MMDX test results (Supplementary Table S3) showed that each 1-SD higher score in the Probable Rejection Model 1 ABMR/Injury score was associated with a 3.92 g (95% CI, 0.22-7.63) higher LVM (p=0.038); 1-SD increase in the Probable Rejection Model 2 ABMR score was associated with a 6.14 g (95% CI, 2.06-10.22) higher LVM (p=0.003) while a higher Probable Rejection Model 2 no rejection/injury transcript was associated with lower LVM (−4.30 g (95% CI, −8.45 to −0.15) per 1 SD; p=0.042). None of the other transcript scores were associated with LVM in the patient comparison. When MMDx results were analyzed as categorical interpretations (Normal, Slightly Abnormal, Abnormal), no association reached statistical significance in either dimension (Supplementary Table S4).

### Transcriptomic exploration

In the independent transcriptomic cohort, patient level remodeling scores represented the spectrum of AGR. 14 patients were classified as AGR and 21 as non-AGR. GSEA analysis using patient-level remodeling score as a continuous risk (and adjusted for biopsy time and donor sex) identified 17 significant Hallmark pathways and 221 significant Gene Ontology pathways (biological processes, molecular function and cellular component) at an FDR <0.05. Increasing remodeling score was associated with enrichment of a prominent interferon associated transcriptional program: (IFN-Alpha -NES: 2.27; FDR< 0.001 and IFN-gamma response -NES:1.57; FDR=0.008). In contrast, lower remodeling scores were associated with enrichment of mitochondrial oxidative-metabolic and biosynthetic programs including oxidative phosphorylation, ATP synthesis-coupled electron transport, cytoplasmic translation, ribosome biogenesis and ribonucleoprotein-complex organization. A higher GR score was also associated with Fc-receptor pathways (Fc receptor mediated stimulatory signaling-NES:2.09; FDR=0.007 and Fc Gamma receptor signaling pathway-NES:1.97; FDR=0.026 pathways). In contrast, the Hallmark mTOR signaling was associated with lower remodeling scores (NES: −1.60; FDR = 0.008). Embryonic programming pathways of developmental growth involved in morphogenesis (NES:1.51; FDR=0.053) and regulation of WNT signaling pathway (NES: 1.48; FDR=0.053) were directionally associated with remodeling score and were close to statistical significance. Similarly, vascular endothelial growth factor receptor (NES:1.74; FDR=0.060) and defense response to virus (NES:1.48; FDR=0.060) GO pathways, while did not meet significance were close to the FDR threshold. Broad canonical allograft-rejection, inflammatory, IL-2/STAT5, IL-6/JAK/STAT3 and TNF/NF-κB pathways were not significantly enriched after FDR correction. (Figure 6, Supplemental Table S5).

**Figure 6.**
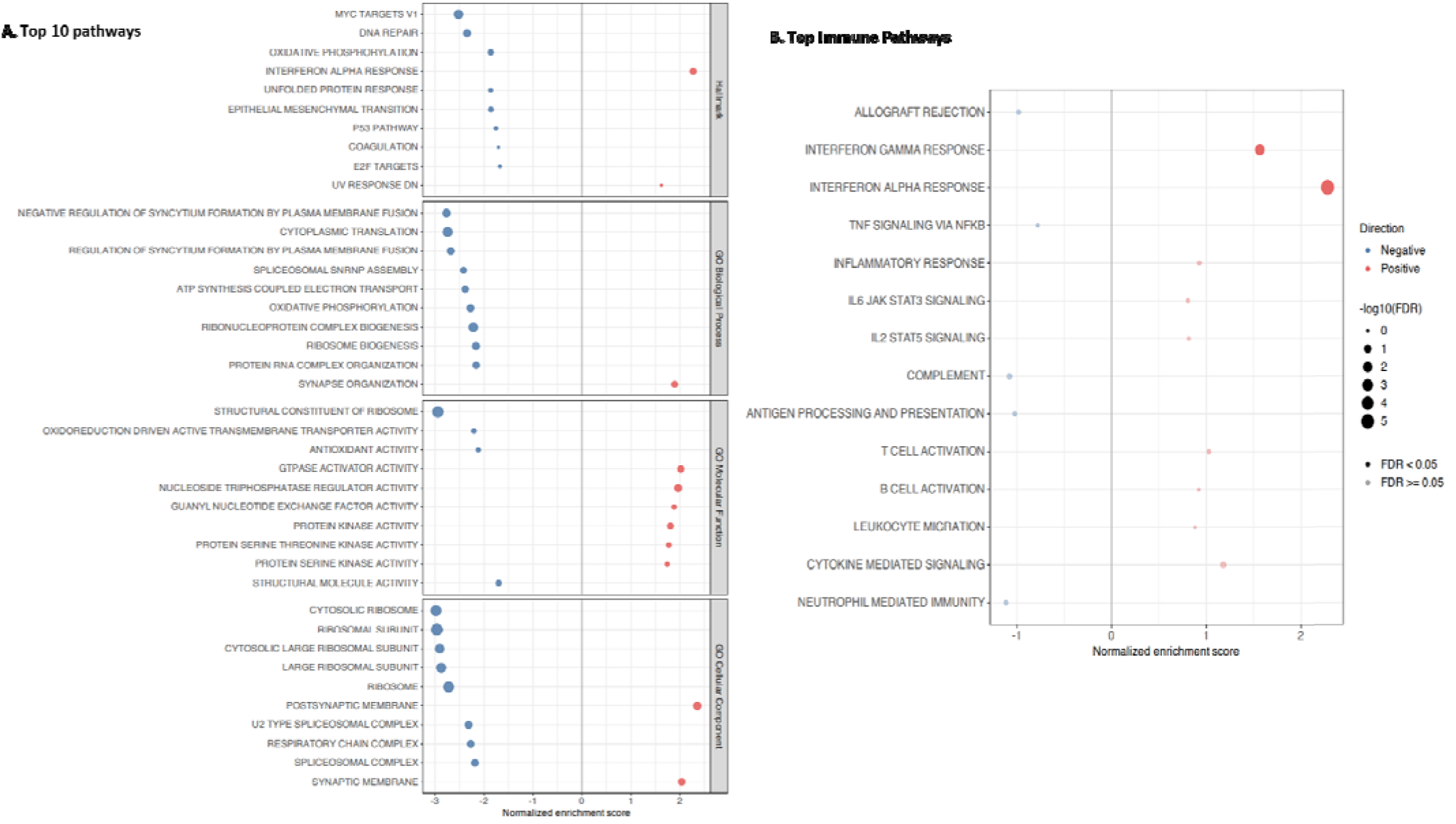
Gene set enrichment analysis (GSEA) in the independent transcriptomic exploration cohort. GSEA of biopsy samples with the remodeling score as a continuous variable, while accounting for repeated biopsies from the same patient and adjusted for time of biopsy and donor sex. Gene set enrichment analysis was performed using all tested genes ranked by the moderated t-statistic. Panel (A) shows top 10 Hallmark and Gene Ontology pathways. Panel (B) shows selected immune pathways. Point size represents −log10(FDR), and gray points indicate FDR ≥0.05. Overall, 17 significant Hallmark pathways and 221 significant Gene Ontology pathways (biological processes, molecular function and cellular component) at an FDR <0.05.

### Agreement between echocardiographic and CMR LVM

Among 236 echocardiogram-CMR pairs obtained within 90 days, LVM measurements were moderately correlated (Pearson r=0.65; ICC, 0.61; 95% CI, 0.53-0.69). Echocardiography overestimated LV mass by a mean of 30.2 g compared with CMR (Supplementary Figure 2).

## DISCUSSION

Long-term outcomes after heart transplant continue to be suboptimal (21) with no advancements in specific therapies that alter the onset and progression of chronic rejection. CAV is typically labeled as the ‘*sine qua non’* of chronic rejection and viewed as a late-onset phenotype. Such a prevailing paradigm limits our understanding of the complex and subtle biological mechanisms that injure the allograft, maintain adaptive and maladaptive remodeling through the lifetime of the allograft, and misses opportunities to discover therapeutic interventions. Goodroe et al showed that 83% of post HT patients in a single-center study had myocardial hypertrophy at 1 year, and LVM >250g was associated (HR, 3.6) with mortality(6). Reliance on LVM assessed at a single time point does not capture the dynamic nature of various insults and creates opportunities for intervention.

Using the TCPRP-heart, an automated, multisource, deeply phenotyped single-center registry (10), we evaluated the relevance of LVM changes through the life of the graft, rather than a binary state of hypertrophy vs no hypertrophy. This distinction is especially important in a transplanted heart as the allograft undergoes substantial and nonuniform adaptation. While early LVM measurements may reflect factors related to the donor and organ procurement process, later measurements may reflect recipient-related factors (metabolic, immune-related, and hemodynamic). At the outset, our data confirms the clinical significance of LVM as a simple imaging biomarker that predicts clinically meaningful outcomes. An association of AGR with GD as an intermediate impact on the heart that ultimately leads to death creates an opportunity to devise strategies to intervene. Apart from the absolute measure of LVM, the dynamic changes (1g/year), reflected in the slope assessment, predicted future GD and future CAV2/3, suggesting the importance of temporal impact on the graft.

Unlike historical histological, myocyte-centric understanding of LVH (22) imaging-based LVM measurement captures mechanisms beyond myocyte afterload (interstitial edema, inflammatory infiltration, and non-cardiomyocyte changes). By using a joint modeling methodology, we were able to factor such impact and found that the most relevant change was a failure to reverse remodel to a lower LVM compared to the group that was able to. Thus, AGR in this context needs to be viewed not as a mere increase of LVM but as a failure of the expected reverse-remodeling response after transplant. This paradigm also reflects the importance of early LVM changes potentially reflecting adaptation from organ procurement and surgical stressors, load and immune adaptation, changes that are new for the heart. Our data does not have the ability to study the impact of recent advances in this space. LVM index analyses were also directionally concordant with non-adjusted measurements, suggesting that the prognostic importance of ongoing remodeling evident on imaging of the heart is independent of the body surface area. Our findings therefore suggest a need to reframe allograft remodeling in the context of imaging, capturing an organ-level change contrary to the classic imagination of load-dependent myocyte hypertrophy.

Several clinical biological domains were associated with LVM. Donor age, body size, recipient sex, and donor-recipient sex pairing were expected baseline nonmodifiable covariates that are known to associate with higher LVM. Hemodynamic load (pulse pressure and systolic pressure) is an important modifiable factor. Specific goals for blood pressure after heart transplant are lacking and our data reconfirms the importance of hypertension with every 10 mm Hg increase in systolic pressure associated with an 8.34 g increase in LVM. Hence, strict control of blood pressure and specific goals are needed. Amongst all the relevant clinical immune-mediated markers, a greater average endothelial DSA-selective transcript activity on MMDx had the greatest average LVM contribution in the between patient comparison, suggesting a possible central biology of endothelial injury driving ventricular remodeling. Our work cannot make conclusive biological underpinnings but is a reconfirmation of studies that hypothesize early endothelial activation to contribute to later adverse outcomes in HT. (23)

Conventional biopsy and MMDx transcripts further supported the importance of graft injury in the within patients’ comparison. LVM was higher during periods associated with acute cellular rejection even at grade 1R. Recent research has suggested that 1R rejection might have more clinical relevance than currently appreciated based on transcript assessment (24) and further studies are needed to address treatment strategies that can mitigate the residual risk mediated by 1R rejection. Also, of note was a positive association of LVM with C4d immunostaining. Higher Probable Rejection Model 1 ABMR/Injury and Probable Rejection Model 2 ABMR activity tracked higher LVM suggesting a continuum of subtle antibody mediated injury. A higher Probable Rejection Model 2 with no rejection/ injury activity was associated with lower LVM defining the importance of quiescence from rejection as a positive impact on the graft. Myocardial injury/stress biomarkers showed similar dynamic relationships, with LV mass increasing alongside NT-proBNP and high-sensitivity troponin T within patients. Together, these findings suggest that serial LVM may integrate graft structure, hemodynamic load, immune-mediated injury, and myocardial stress rather than reflecting one isolated mechanism. Prior mechanistic studies have similarly suggested that post-transplant hypertrophy is not merely hemodynamic mediated. Stetson et al.(25) identified persistent myocardial tumor necrosis factor-α expression in association with cardiomyocyte hypertrophy and fibrosis and a more recent study suggested that everolimus-based treatment, may influence LVM(26). Such findings extend the possibilities of intervenable strategies to target biologically modifiable mechanisms. Our data did not show any association of harm or protection of immunosuppression levels of tacrolimus or proliferation signal inhibitors. Of note, our center primarily utilized tacrolimus-based regimen during the study period.

The independent transcriptomic cohort further validates the importance of early changes in the graft providing insights from surveillance biopsies early after HT. Biopsies from patients with a higher graft remodeling score were enriched with an interferon-responsive state rather than a generalized transcriptional signature of conventional rejection. A recent study using spatial transcriptomics supported an interferon responsive molecular state centered around macrophages.(27) Several vascular and endothelial pathways (including developmental cell fate pathways) were also directionally enriched with increasing remodeling scores. Together with the interferon response signature, these findings raise the hypothesis that immune associated endothelial activation may contribute to subsequent AGR. The enrichment of Fc receptor pathways validates the clinical relevance of antibody mechanisms portending adverse survival and supports the observation that Fc gamma inhibitory strategy was beneficial to prevent chronic rejection in a mouse model. (28) A positive DSA (presence of class I or II at MFI>2,000) did not show a significant association in our study, raising the possibility of role of non-DSA, non-HLA and cardiac antibodies (not studied in our cohort). Similarly the hallmark mTOR pathway validates the RADTAC study that showed addition of a low dose mTOR inhibitor attenuated hypertrophy in the first year after HT.(29) Conversely, lower remodeling scores were associated with relative enrichment of mitochondrial energy-production and cellular biosynthesis programs. These findings suggest that the adverse remodeling phenotype may reflect early immune activation and graft injury, whereas more favorable remodeling may be associated with preservation of myocardial structural and metabolic processes with mitochondrial reserve. Because these analyses were performed in bulk tissue, the observed difference may reflect changes in cellular composition, transcriptional state within resident cells or both and should hence be considered hypothesis generating rather than evidence of a specific causal mechanism.

The exploratory CAV analysis from angiograms performed after 4 years from transplant, extends the broader concept of a continuum of myocardial injury response reflected in LVM changes leading ultimately to the end stage of chronic rejection. A more positive LVM slope, but not a onetime LVM, was associated with subsequent angiographic CAV grade 2 or 3. Such biological plausibility of myocardial vascular changes preceding long-term adverse outcomes is backed by previous studies showing a high burden of stenotic micro-vasculopathy in myocardial biopsies (within the first year) associated with long-term mortality. (23) The corresponding association between time-updated CAV and mortality confirms the adverse prognosis of established angiographic disease.

Our study has various limitations apart from being a single center study. We were unable to obtain LVM in the donor prior to procurement (due to limited access to such reports which do not populate the EHR) limiting our ability to delineate the impact of organ procurement vs donor derived risk. The LVM calculation used NLP based algorithm to extract the relevant variables from the report that can limit its translatability across centers with limited quality control. Our center maintains high-quality control in the cardiovascular imaging department and hence has reliability of measurements. Such a claim was validated by a fair correlation with MRI based LVM. We chose to utilize echocardiogram based LVM measurements due to its ability to provide an easy, cost effective and generalizable technology. Current advancements in artificial intelligence based echocardiography software that can measure LVM automatically can help overcome reliance on manual measurements.(30)

Despite these limitations, this study has several important strengths, including the large number of serial echocardiographic measurements, use of transplantation as a common time origin, exclusion of measurements immediately preceding outcomes, and application of joint models that accounted for the interdependence between longitudinal LVM and clinical events.

Joint modeling was a central strength to our analysis. A conventional mixed-effects model can describe LVM over time, while a separate Cox model can estimate event risk, but neither approach alone directly relates the evolving LVM trajectory to the timing of GD or death(14, 31, 32). Prior approaches of using a Cox model based on one LVM value at a specific time interval discard most of the longitudinal information. Similarly, directly using predetermined intercept and slope for each patient in a Cox model reduces each patient’s trajectory to a starting value and a single average rate of change. In contrast, joint modeling incorporates all available measurements and accounts for irregular imaging intervals, nonlinear changes, measurement variability, and uncertainty. This approach is particularly important in this context because GD or death may stop further imaging(33, 34). The availability of a deeply phenotyped automated registry also enabled evaluation of structural, hemodynamic, immunologic, molecular, and biochemical correlations.

## Conclusion

GR post heart transplant is an easily available imaging biomarker reflecting the complex changes the transplanted heart undergoes adapting to a hostile (immunological and non-immunological) environment. The association of AGR with clinically meaningful adverse outcomes (GD, CAV and death) provides an opportunity to identify patients at risk and create strategies to intervene. Apart from clinically modifiable factors, the presence of distinct biological states in the transcriptomic analysis could provide opportunities to discover treatable targets. Further multicenter collaboration to validate, replicate the data infrastructure and invest in exploring biological mechanisms could lead to therapies to protect the graft beyond just an antirejection strategy after HT.

## Data Availability

The data used in this study are institutional data protected under institutional review board and HIPAA restrictions and cannot be shared publicly.

## Author Contributions

AB and KP contributed to study conception and design. KP contributed to data acquisition, data curation, statistical analysis, and interpretation of data. KP drafted the manuscript, and AB revised and updated it. RR supported the transcriptomics sub-study in sample collection, processing, shipping, and review of the manuscript. TP and RG provided transcriptomic assessment, analysis, and manuscript review. SAK, TE, GTA, AG, and RG critically reviewed and revised the manuscript for important intellectual content. All authors approved the final version.

## Abbreviations

ABMR = antibody-mediated rejection, ACR = acute cellular rejection, AGR = adverse graft remodeling, CAV = cardiac allograft vasculopathy, CMR = cardiac magnetic resonance, cPRA = calculated panel-reactive antibody, CrI = credible interval, CI= Confidence Interval, DBD = donation after brain death, DCD = donation after circulatory death, DSA = donor-specific antibody, DSAST = DSA-selective transcripts, eDSAST = endothelial DSA-selective transcripts, EHR = electronic health record, FDR = false discovery rate, GD = graft dysfunction, GO = Gene Ontology, GR = graft remodeling, GSEA = gene set enrichment analysis, HT = heart transplantation, HT1 = heart transcripts, ICC = intraclass correlation coefficient, IVSD = interventricular septal thickness in diastole, LVH = left ventricular hypertrophy, LVIDD = left ventricular internal diameter in diastole, LVM = left ventricular mass, MFI = mean fluorescence intensity, MMDx = Molecular Microscope Diagnostic System, MSigDB = Molecular Signatures Database, NES = normalized enrichment score, NLP = natural language processing, non-AGR = no adverse graft remodeling, NRI = non-rejection injury, PWD = posterior wall thickness in diastole, TCB = T cell burden transcripts, TCMR = T cell-mediated rejection, TCPR = Transplant Center Precision Registry

---

**Supplemental Figure 1.**
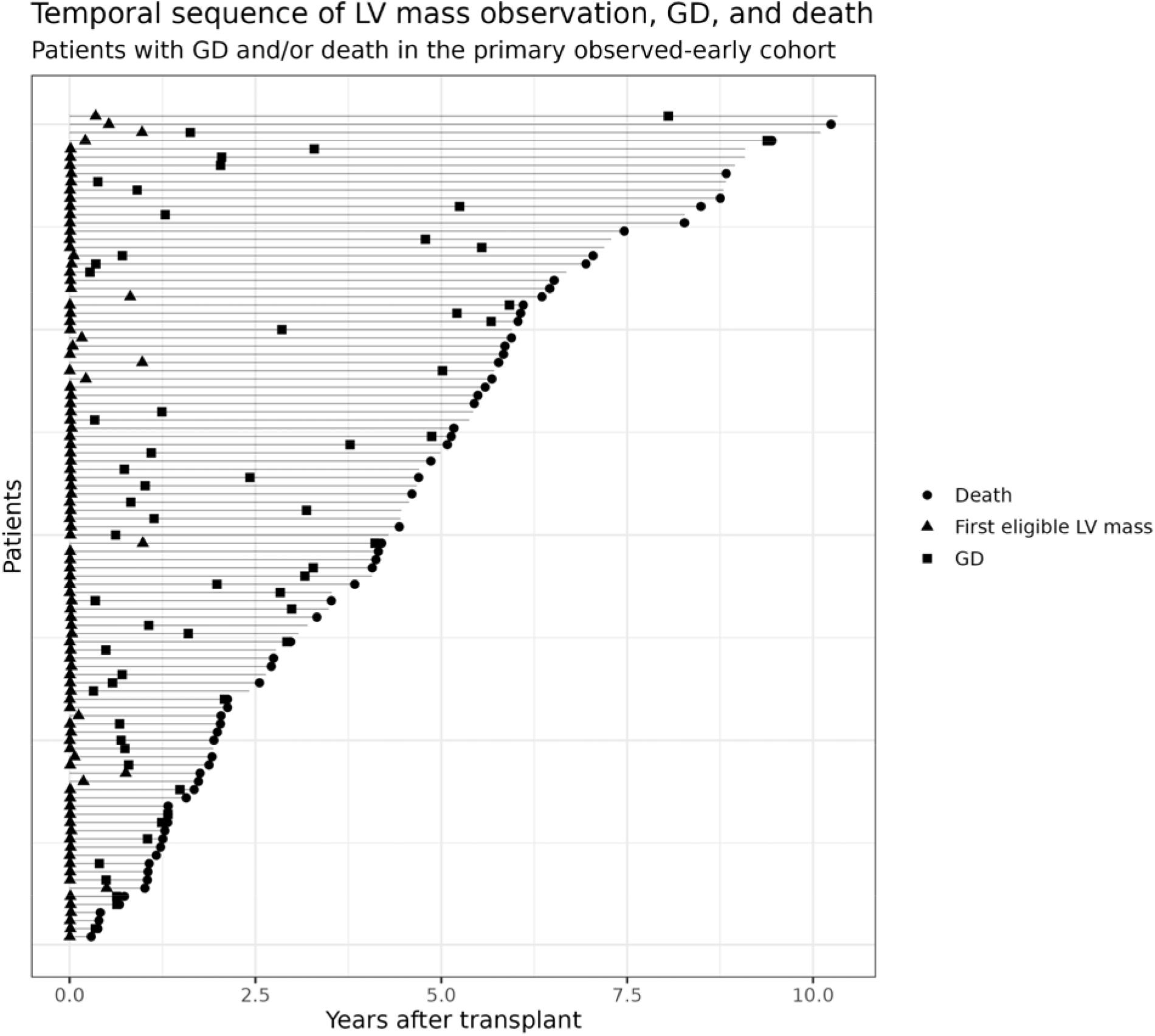
T**e**mporal **sequence of LV mass observation, graft dysfunction, and death.** Each horizontal line represents a patient in the primary cohort who experienced graft dysfunction, death, or both. Triangles indicate the first eligible LV mass measurement, squares indicate the first graft-dysfunction event, and circles indicate death. Patients ordered by descending order of followup time.

**Supplemental Figure 2.**
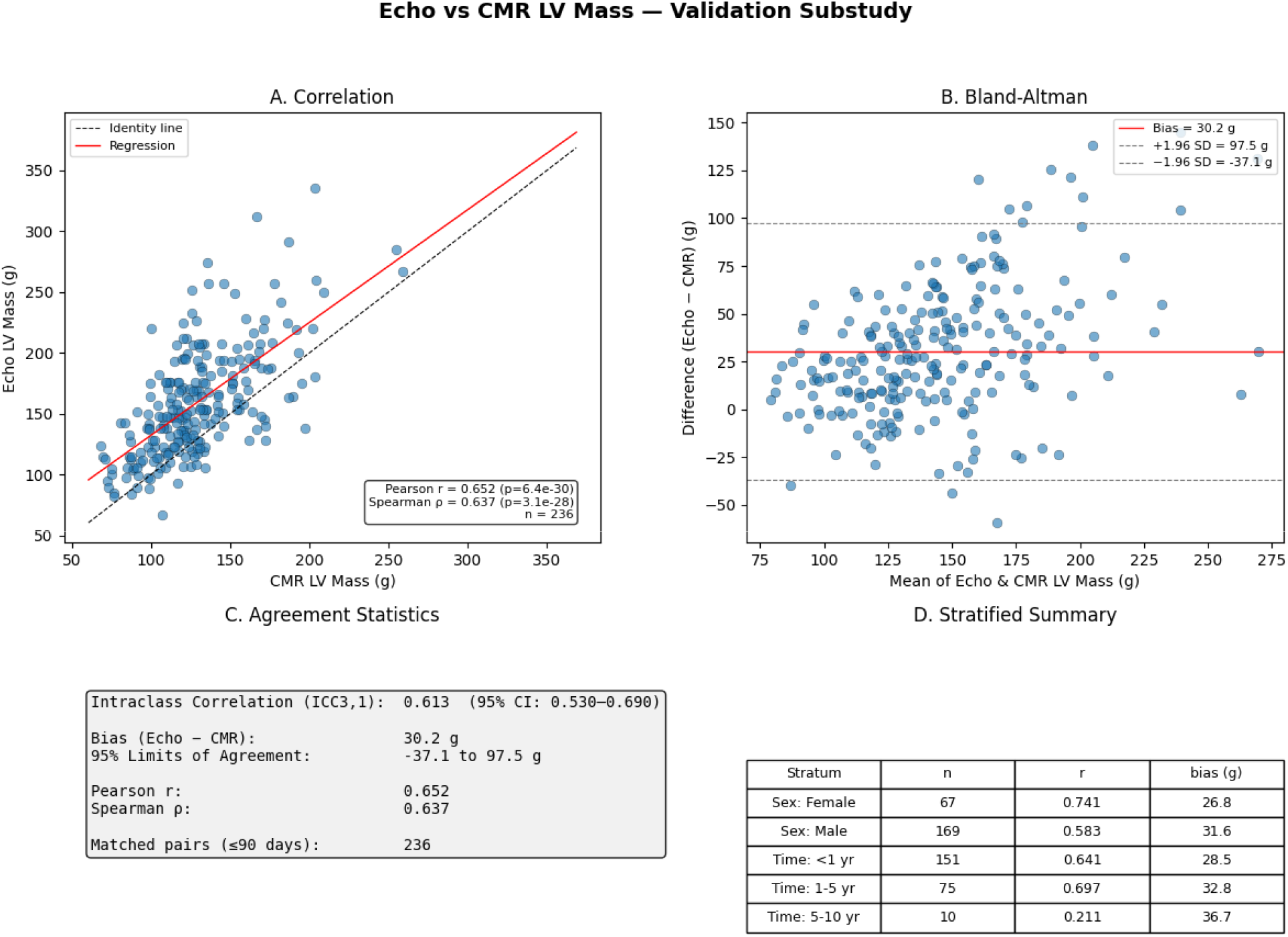
A**g**reement **between echocardiographic and cardiac magnetic resonance–derived left ventricular mass.** A total of 236 echocardiogram–cardiac magnetic resonance pairs obtained within 90 days were included. **A**, Correlation between echocardiographic and CMR-derived LV mass. The dashed line represents the line of identity and the solid line represents the fitted linear regression. **B**, Bland–Altman plot showing the difference between echocardiographic and CMR-derived LV mass against their mean. The solid horizontal line indicates the mean bias and dashed lines indicate the 95% limits of agreement. **C**, Summary agreement statistics. Echocardiographic and CMR-derived LV mass showed moderate agreement (intraclass correlation coefficient, 0.613; 95% CI, 0.530–0.690) and moderate correlation (Pearson *r*=0.652; Spearman ρ=0.637). Echocardiography overestimated LV mass by a mean of 30.2 g, with 95% limits of agreement from −37.1 to 97.5 g. **D**, Stratified correlations and mean biases according to recipient sex and time after transplantation. CMR indicates cardiac magnetic resonance; ICC, intraclass correlation coefficient; LV, left ventricular.

