## Supplementary Tables S1-S4 for "Adverse Graft Remodeling Reflects Dynamic Allograft Stress and Predicts Adverse Outcomes After Heart Transplantation"

**Supplementary Tables. Within-patient comparisons**

Supplementary Tables S1-S4 report WITHIN-PATIENT associations: each patient serves as his or her own control, and the estimate compares periods when that patient's exposure was above or below that same patient's own average. They answer the question, "When this variable rises above a patient's usual level, does that patient's left ventricular mass rise with it?" Because the comparison is made inside a single patient, these estimates are not confounded by fixed differences between patients such as body size, sex, or donor characteristics. Only time-varying exposures can be decomposed into a within-patient component, so fixed donor, recipient, and baseline immunologic characteristics do not appear in these tables. All between-patient results are shown in Table 2 of the main manuscript.

**Supplementary Table S1. Within-patient associations of hemodynamic, laboratory, cardiac biomarker, metabolic, lipid, and immunosuppressant measures with longitudinal left ventricular mass**

*Estimates reflect deviation of the exposure from that patient's own average value.*

| **Variable** | **Scale / contrast** | **Observations** | **Patients** | **Adjusted difference in LV mass, g (95% CI)** | **p value** |
| --- | --- | --- | --- | --- | --- |
| Pulse pressure | per 10 mmHg | 4,221 | 425 | **1.42 (0.53 to 2.32)** | **0.002** |
| Systolic BP | per 10 mmHg | 4,221 | 425 | **0.84 (0.19 to 1.48)** | **0.011** |
| Mean arterial pressure | per 10 mmHg | 4,221 | 425 | 0.69 (-0.23 to 1.62) | 0.141 |
| Sodium | per 1 mmol/L | 5,523 | 439 | -0.21 (-0.48 to 0.06) | 0.129 |
| Potassium | per 1 mmol/L | 5,530 | 439 | **2.01 (0.14 to 3.88)** | **0.035** |
| Chloride | per 1 mmol/L | 5,523 | 439 | 0.09 (-0.15 to 0.34) | 0.465 |
| CO2 | per 1 mmol/L | 5,523 | 439 | -0.04 (-0.34 to 0.27) | 0.822 |
| Anion gap | per 1 mmol/L | 5,514 | 439 | **-0.56 (-0.91 to -0.21)** | **0.002** |
| BUN | per 10 mg/dL | 5,523 | 439 | **0.69 (0.03 to 1.35)** | **0.040** |
| Creatinine | per 1 mg/dL | 5,523 | 439 | -0.10 (-1.38 to 1.18) | 0.877 |
| Glucose | per 10 mg/dL | 5,523 | 439 | -0.15 (-0.36 to 0.05) | 0.148 |
| Calcium | per 1 mg/dL | 5,523 | 439 | **-4.15 (-5.57 to -2.74)** | **<0.001** |
| Total protein | per 1 g/dL | 5,495 | 439 | **-3.21 (-4.36 to -2.07)** | **<0.001** |
| Albumin | per 1 g/dL | 5,494 | 439 | **-4.94 (-6.51 to -3.37)** | **<0.001** |
| Albumin/globulin ratio | per 1 unit | 4,649 | 439 | **-5.79 (-9.07 to -2.52)** | **<0.001** |
| Alkaline phosphatase | per 10 U/L | 5,494 | 439 | 0.01 (-0.11 to 0.13) | 0.892 |
| AST | per 10 U/L | 5,494 | 439 | -0.07 (-0.16 to 0.03) | 0.161 |
| ALT | per 10 U/L | 5,495 | 439 | -0.08 (-0.18 to 0.02) | 0.122 |
| Total bilirubin | per 1 mg/dL | 5,495 | 439 | **-2.81 (-4.10 to -1.51)** | **<0.001** |
| Tacrolimus trough | per 1 ng/mL | 2,595 | 352 | 0.06 (-0.25 to 0.37) | 0.708 |
| Sirolimus trough | per 1 ng/mL | 98 | 26 | -1.59 (-4.66 to 1.47) | 0.303 |
| NT-proBNP | per doubling | 986 | 238 | **4.86 (3.44 to 6.29)** | **<0.001** |
| High-sensitivity troponin T | per doubling | 1,590 | 284 | **1.77 (0.90 to 2.65)** | **<0.001** |
| Creatine kinase | per doubling | 668 | 144 | -2.36 (-5.40 to 0.68) | 0.128 |
| Total cholesterol | per 10 mg/dL | 3,437 | 413 | 0.32 (-0.04 to 0.68) | 0.082 |
| Triglycerides | per doubling | 3,565 | 417 | -1.18 (-3.23 to 0.86) | 0.257 |
| HDL cholesterol | per 10 mg/dL | 3,043 | 405 | 0.91 (-0.05 to 1.86) | 0.063 |
| LDL cholesterol | per 10 mg/dL | 3,026 | 405 | 0.43 (-0.08 to 0.95) | 0.101 |
| Hemoglobin A1c | per 1 percentage point | 3,341 | 408 | **-1.78 (-3.08 to -0.48)** | **0.007** |

Blood pressure was matched to the closest preceding value within 180 days. Routine laboratory values, cardiac biomarkers, and immunosuppressant trough concentrations were matched within 90 days. NT-proBNP, high-sensitivity troponin T, creatine kinase, and triglycerides were modeled using log2(value + 1); estimates are interpreted as the approximate difference in LV mass per doubling of the transformed biomarker.

*Values in bold are statistically significant at p<0.05. p values are unadjusted for multiple comparisons.*

**Supplementary Table S2. Within-patient associations of conventional biopsy rejection measures with longitudinal left ventricular mass**

*Each patient serves as his or her own control; estimates compare periods when the exposure was above or below that same patient's average.*

| **Variable** | **Scale / contrast** | **Adjusted difference in LV mass, g (95% CI)** | **p value** | **LV-mass observations** | **Patients** | **Patients with within-person variation** |
| --- | --- | --- | --- | --- | --- | --- |
| ACR grade >=1R | Closest preceding biopsy: ACR grade >=1R vs <1R | **3.60 (1.50 to 5.69)** | **<0.001** | 5,533 | 439 | 371 |
| C4d immunostain-based AMR proxy | per 10-percentage-point increase | **1.56 (0.53 to 2.60)** | **0.003** | 5,461 | 439 | 318 |

The closest preceding biopsy within 180 days was used; same-day biopsies were excluded. When no biopsy was available, rejection exposures were coded as zero. If a biopsy was present but the relevant pathology result was missing, the exposure was treated as missing.

*Values in bold are statistically significant at p<0.05. p values are unadjusted for multiple comparisons.*

**Supplementary Table S3. Within-patient associations of continuous MMDx transcript scores with longitudinal left ventricular mass**

*Exploratory subgroup analysis of 497 paired LV-mass/MMDx observations from 80 patients.*

| **Variable** | **Scale / contrast** | **Adjusted difference in LV mass, g (95% CI)** | **p value** | **Observations** | **Patients** | **Patients with within-person variation** |
| --- | --- | --- | --- | --- | --- | --- |
| Probable Rejection Model 1 TCMR/Injury | per 1 SD higher score | -0.13 (-4.02 to 3.75) | 0.947 | 497 | 80 | 58 |
| Probable Rejection Model 1 NRI | per 1 SD higher score | -2.32 (-6.11 to 1.46) | 0.228 | 497 | 80 | 53 |
| Probable Rejection Model 1 ABMR/Injury | per 1 SD higher score | **3.92 (0.22 to 7.63)** | **0.038** | 497 | 80 | 58 |
| Probable Rejection Model 2 TCMR | per 1 SD higher score | -1.66 (-5.59 to 2.28) | 0.409 | 497 | 80 | 58 |
| Probable Rejection Model 2 NRI | per 1 SD higher score | **-4.30 (-8.45 to -0.15)** | **0.042** | 497 | 80 | 49 |
| Probable Rejection Model 2 ABMR | per 1 SD higher score | **6.14 (2.06 to 10.22)** | **0.003** | 497 | 80 | 56 |
| Enzyme (ADAMDEC1) | per 1 SD higher score | 2.79 (-1.37 to 6.96) | 0.188 | 497 | 80 | 58 |
| Checkpoint (CTLA4) | per 1 SD higher score | 0.84 (-3.06 to 4.73) | 0.674 | 497 | 80 | 58 |
| Cytokine (CXCL13) | per 1 SD higher score | -0.78 (-5.31 to 3.75) | 0.736 | 497 | 80 | 58 |
| DSA-selective (DSAST) | per 1 SD higher score | 0.24 (-4.84 to 5.32) | 0.927 | 497 | 80 | 58 |
| IFNG inducible (GRIT) | per 1 SD higher score | 3.95 (-0.83 to 8.74) | 0.105 | 497 | 80 | 58 |
| Heart transcripts (HT1) | per 1 SD higher score | -3.47 (-7.85 to 0.92) | 0.121 | 497 | 80 | 58 |
| Interferon gamma (IFNG) | per 1 SD higher score | 2.56 (-1.26 to 6.37) | 0.188 | 497 | 80 | 58 |
| Injury transcripts (IRRAT) | per 1 SD higher score | 0.82 (-3.30 to 4.95) | 0.695 | 497 | 80 | 58 |
| NK cell burden (NKB) | per 1 SD higher score | 2.97 (-1.57 to 7.50) | 0.199 | 497 | 80 | 58 |
| PC1 | per 1 SD higher score | 2.95 (-1.77 to 7.67) | 0.220 | 497 | 80 | 58 |
| PC2 | per 1 SD higher score | -1.45 (-5.37 to 2.48) | 0.469 | 497 | 80 | 58 |
| PC3 | per 1 SD higher score | 1.24 (-3.17 to 5.65) | 0.582 | 497 | 80 | 58 |
| Cytotoxic T cell transcripts (QCAT) | per 1 SD higher score | 2.59 (-2.07 to 7.25) | 0.275 | 497 | 80 | 58 |
| Macrophage transcripts (QCMAT) | per 1 SD higher score | 2.71 (-1.89 to 7.30) | 0.247 | 497 | 80 | 58 |
| Rejection Model 1 NRI | per 1 SD higher score | -2.43 (-7.23 to 2.37) | 0.321 | 497 | 80 | 58 |
| Rejection Model 1 TCMR/Injury | per 1 SD higher score | 2.30 (-1.77 to 6.38) | 0.267 | 497 | 80 | 52 |
| Rejection Model 1 ABMR/Injury | per 1 SD higher score | 0.98 (-3.59 to 5.55) | 0.673 | 497 | 80 | 55 |
| Rejection Model 2 NRI | per 1 SD higher score | -3.03 (-7.87 to 1.80) | 0.218 | 497 | 80 | 58 |
| Rejection Model 2 TCMR | per 1 SD higher score | -0.43 (-4.01 to 3.15) | 0.813 | 497 | 80 | 42 |
| Rejection Model 2 ABMR | per 1 SD higher score | 1.34 (-3.55 to 6.23) | 0.590 | 497 | 80 | 55 |
| Rejection Model 2 Injury | per 1 SD higher score | 3.17 (-1.15 to 7.48) | 0.150 | 497 | 80 | 46 |
| ROBO4 | per 1 SD higher score | -1.86 (-6.67 to 2.95) | 0.448 | 497 | 80 | 58 |
| T cell burden (TCB) | per 1 SD higher score | 2.14 (-2.58 to 6.86) | 0.374 | 497 | 80 | 58 |
| Endothelial DSA-selective (eDSAST) | per 1 SD higher score | -3.28 (-7.35 to 0.80) | 0.115 | 497 | 80 | 58 |

Continuous MMDx scores were standardized; estimates represent the difference in LV mass per 1-SD higher score relative to that patient's own average. The closest preceding MMDx assessment within 180 days was used, and same-day assessments were excluded.

*Values in bold are statistically significant at p<0.05. p values are unadjusted for multiple comparisons*

**Supplementary Table S4. Within-patient associations of categorical MMDx interpretations with longitudinal left ventricular mass**

*The current interpretation is compared with Normal relative to that patient's usual pattern.*

| **Variable** | **Scale / contrast** | **Adjusted difference in LV mass, g (95% CI)** | **p value** | **Observations** | **Patients** | **Patients with category change** |
| --- | --- | --- | --- | --- | --- | --- |
| Enzyme (ADAMDEC1), Slightly Abnormal | Slightly Abnormal vs Normal | 1.96 (-6.67 to 10.58) | 0.656 | 497 | 80 | 38 |
| Enzyme (ADAMDEC1), Abnormal | Abnormal vs Normal | 5.74 (-11.46 to 22.94) | 0.512 | 497 | 80 | 38 |
| Checkpoint (CTLA4), Slightly Abnormal | Slightly Abnormal vs Normal | -0.37 (-11.59 to 10.86) | 0.949 | 497 | 80 | 22 |
| Checkpoint (CTLA4), Abnormal | Abnormal vs Normal | 0.33 (-21.32 to 21.99) | 0.976 | 497 | 80 | 22 |
| Cytokine (CXCL13), Slightly Abnormal | Slightly Abnormal vs Normal | 2.13 (-10.52 to 14.77) | 0.741 | 497 | 80 | 16 |
| Cytokine (CXCL13), Abnormal | Abnormal vs Normal | -3.15 (-29.05 to 22.75) | 0.811 | 497 | 80 | 16 |
| DSA-selective (DSAST), Slightly Abnormal | Slightly Abnormal vs Normal | -10.17 (-23.37 to 3.03) | 0.131 | 497 | 80 | 24 |
| DSA-selective (DSAST), Abnormal | Abnormal vs Normal | 10.06 (-3.63 to 23.75) | 0.149 | 497 | 80 | 24 |
| IFNG inducible (GRIT), Slightly Abnormal | Slightly Abnormal vs Normal | 0.77 (-12.67 to 14.21) | 0.910 | 497 | 80 | 29 |
| IFNG inducible (GRIT), Abnormal | Abnormal vs Normal | 1.08 (-9.95 to 12.10) | 0.848 | 497 | 80 | 29 |
| Heart transcripts (HT1), Abnormal | Abnormal vs Normal | 8.12 (-0.99 to 17.23) | 0.080 | 497 | 80 | 30 |
| Interferon gamma (IFNG), Slightly Abnormal | Slightly Abnormal vs Normal | -3.63 (-15.17 to 7.91) | 0.537 | 497 | 80 | 27 |
| Interferon gamma (IFNG), Abnormal | Abnormal vs Normal | 4.06 (-10.53 to 18.65) | 0.585 | 497 | 80 | 27 |
| Injury transcripts (IRRAT), Slightly Abnormal | Slightly Abnormal vs Normal | 0.43 (-9.68 to 10.53) | 0.934 | 497 | 80 | 37 |
| Injury transcripts (IRRAT), Abnormal | Abnormal vs Normal | 3.32 (-7.71 to 14.35) | 0.555 | 497 | 80 | 37 |
| NK cell burden (NKB), Slightly Abnormal | Slightly Abnormal vs Normal | 4.96 (-8.80 to 18.71) | 0.479 | 497 | 80 | 24 |
| NK cell burden (NKB), Abnormal | Abnormal vs Normal | 6.37 (-7.00 to 19.75) | 0.349 | 497 | 80 | 24 |
| Cytotoxic T cell transcripts (QCAT), Slightly Abnormal | Slightly Abnormal vs Normal | 5.75 (-6.42 to 17.91) | 0.354 | 497 | 80 | 26 |
| Cytotoxic T cell transcripts (QCAT), Abnormal | Abnormal vs Normal | 4.11 (-9.11 to 17.33) | 0.542 | 497 | 80 | 26 |
| Macrophage transcripts (QCMAT), Slightly Abnormal | Slightly Abnormal vs Normal | -4.99 (-16.98 to 7.01) | 0.414 | 497 | 80 | 30 |
| Macrophage transcripts (QCMAT), Abnormal | Abnormal vs Normal | 6.35 (-5.56 to 18.25) | 0.295 | 497 | 80 | 30 |
| ROBO4, Slightly Abnormal | Slightly Abnormal vs Normal | 0.49 (-10.13 to 11.10) | 0.928 | 497 | 80 | 27 |
| ROBO4, Abnormal | Abnormal vs Normal | 4.49 (-13.06 to 22.04) | 0.616 | 497 | 80 | 27 |
| Injury cluster (S4), Slightly Abnormal | Slightly Abnormal vs Normal | -9.36 (-23.63 to 4.91) | 0.198 | 497 | 80 | 25 |
| Injury cluster (S4), Abnormal | Abnormal vs Normal | 5.10 (-7.35 to 17.54) | 0.421 | 497 | 80 | 25 |
| T cell burden (TCB), Slightly Abnormal | Slightly Abnormal vs Normal | -4.65 (-17.19 to 7.90) | 0.467 | 497 | 80 | 25 |
| T cell burden (TCB), Abnormal | Abnormal vs Normal | 3.87 (-9.83 to 17.56) | 0.579 | 497 | 80 | 25 |
| Endothelial DSA-selective (eDSAST), Slightly Abnormal | Slightly Abnormal vs Normal | -3.21 (-15.85 to 9.43) | 0.618 | 497 | 80 | 21 |
| Endothelial DSA-selective (eDSAST), Abnormal | Abnormal vs Normal | -9.24 (-32.81 to 14.32) | 0.441 | 497 | 80 | 21 |

Categorical interpretations were modeled as Normal, Slightly Abnormal, or Abnormal.

*Values in bold are statistically significant at p<0.05. p values are unadjusted for multiple comparisons.*

Abbreviations: ABMR, antibody-mediated rejection; ACR, acute cellular rejection; ALT, alanine aminotransferase; AMR, antibody-mediated rejection; AST, aspartate aminotransferase; BMI, body mass index; BP, blood pressure; BUN, blood urea nitrogen; CI, confidence interval; CMV, cytomegalovirus; CO2, carbon dioxide/bicarbonate; cPRA, calculated panel-reactive antibody; DSA, donor-specific antibody; EBV, Epstein-Barr virus; eDSAST, endothelial DSA-selective transcripts; HDL, high-density lipoprotein; LDL, low-density lipoprotein; LV, left ventricular; MMDx, Molecular Microscope Diagnostic System; NRI, non-rejection injury; NT-proBNP, N-terminal pro-B-type natriuretic peptide; SD, standard deviation; TCMR, T cell-mediated rejection.
