## Supplementary Methods for "Adverse Graft Remodeling Reflects Dynamic Allograft Stress and Predicts Adverse Outcomes After Heart Transplantation"

### 1. Study design, data source, and time origin

We conducted a longitudinal observational study of heart transplant recipients enrolled in the J.C. Walter Jr. Transplant Center Precision Registry for Heart. The registry integrates structured electronic health record data with selected variables extracted from unstructured clinical reports using natural language processing.

The date of heart transplantation was used as the common time origin. Echocardiographic measurements, clinical events, catheterization procedures, and censoring times were calculated as days after transplantation and converted to years by dividing by 365.25. Patients could contribute multiple eligible LV mass measurements during follow-up.

The primary analyses evaluated whether the underlying level and rate of change of LV mass were associated with subsequent graft dysfunction and death. An exploratory joint model evaluated cardiac allograft vasculopathy. Additional time-updated analyses examined the temporal associations of graft dysfunction and CAV with mortality.

### 2. Echocardiographic measurements

#### 2.1 Extraction of echocardiographic variables

Left ventricular ejection fraction, left ventricular internal dimension at end diastole, interventricular septal thickness at end diastole, and posterior-wall thickness at end diastole were extracted from clinical echocardiographic reports.

The source measurements were obtained by certified cardiac sonographers within a single health system according to routine clinical standards. Measurements recorded in millimeters were converted to centimeters before calculation of LV mass.

#### 2.2 LV mass calculation

LV mass was calculated using the American Society of Echocardiography–corrected Devereux equation:

LV mass (g)=0.8[1.04{(LVIDD+IVSD+PWD)^3^−(LVIDD)^3^}]+0.6

LV mass index was calculated by dividing LV mass by recipient body surface area.

Body surface area was obtained from the corresponding echocardiographic report. Raw LV mass was the primary longitudinal marker. LV mass index was used only in analyses specifically identified as indexed sensitivity analyses.

#### 2.3 Scaling in joint models

LV mass was divided by 10 during joint-model estimation for numerical scaling. Current-value associations are therefore reported per 10-g higher underlying LV mass.

The fitted slope association was initially expressed per 10-g/year greater LV mass slope. For presentation per 1 g/year, the hazard ratio and credible interval limits were converted by taking their tenth roots.

### 3. Cardiac magnetic resonance validation

Among patients who underwent clinically indicated cardiac magnetic resonance imaging, CMR-derived LV mass was compared with the nearest echocardiographic LV mass measurement obtained within 90 days.

Agreement was evaluated using the Pearson correlation coefficient, intraclass correlation coefficient, and Bland–Altman analysis. Each eligible echocardiogram–CMR pair contributed one paired comparison.

### 4. Clinical outcome definitions

#### 4.1 Graft dysfunction

Graft dysfunction was defined from serial echocardiographic LVEF measurements. For each echocardiogram, the current LVEF was compared with the most recent preceding nonmissing LVEF.

An echocardiogram met the graft-dysfunction definition when either:

1. the current LVEF was below 50%; or
2. the current LVEF was no greater than 75% of the preceding value, corresponding to a relative decline of at least 25%.

The patient-level graft-dysfunction date was the date of the first echocardiogram meeting either criterion. Patients who did not develop graft dysfunction were censored at the date of their last available echocardiogram.

Patients whose first graft-dysfunction event occurred on or before post-transplant day 90 were excluded from the primary LV mass remodeling cohort to avoid capturing primary graft dysfunction.

#### 4.2 Mortality

Mortality was ascertained from institutional records and United Network for Organ Sharing documentation. Time to death or censoring was measured from the date of transplantation. Patients without a recorded death were censored at their last documented registry follow-up.

#### 4.3 Cardiac allograft vasculopathy

All available coronary angiograms were reviewed by a single experienced transplant cardiologist. CAV was graded as 0, 1, 2, or 3 according to International Society for Heart and Lung Transplantation criteria, modified to exclude graft dysfunction from the grade 3 definition. CAV classification was therefore based on angiographic findings alone.

The primary CAV endpoint for the LV mass joint model was the first observed angiogram showing CAV grade 2 or 3. Patients who did not develop CAV grade 2 or 3 were censored at their last available angiogram. CAV grade 2 or 3 recorded within the first 90 days after transplantation was excluded from the joint-model analysis.

### 5. Construction of analytic cohorts

#### 5.1 Primary LV mass remodeling cohort

The primary cohort required all of the following:

1. no graft-dysfunction event on or before post-transplant day 90;
2. at least two usable post-transplant LV mass measurements; and
3. at least one usable LV mass measurement within the first 365.25 days after transplantation.

This yielded 441 eligible patients, of whom 439 and 436 contributed to the graft-dysfunction and mortality joint models respectively after the outcome-specific restrictions described in Section 5.2.

#### 5.2 Event-specific longitudinal datasets

Separate longitudinal datasets were constructed for graft dysfunction and mortality.

For patients who subsequently developed graft dysfunction, eligible LV mass measurements were required to occur before the graft-dysfunction event and at least 91 days before the event. Measurements obtained after graft dysfunction or during the 90 days immediately preceding graft dysfunction were excluded.

For patients who died, eligible LV mass measurements were required to occur before death and at least 91 days before death. For patients without a death event, measurements after the censoring time were excluded. The graft-dysfunction and mortality datasets were constructed from the same 441 patients meeting the primary cohort criteria, but the outcome-specific lead-time restrictions were applied before the two-measurement minimum was re-imposed. This produced non-identical final samples in each direction. Five recipients who died 29 to 93 days after transplantation had no LV mass measurement obtained at least 91 days before death and could not contribute to the mortality joint model. Two recipients who developed graft dysfunction after day 90 retained fewer than two measurements once the graft-dysfunction lead-time restriction was applied and could not contribute to the graft-dysfunction joint model, but retained eligible measurements under the mortality restriction. The graft-dysfunction joint model therefore included 439 patients and 60 events, and the mortality joint model included 436 patients and 71 deaths. Because survival times and outcome status were fully ascertained for all 439 primary-cohort patients, descriptive, Kaplan–Meier, and time-updated Cox analyses of mortality used the full cohort of 439 patients and 75 deaths.

#### 5.3 Exploratory CAV cohort

Because clinically significant vasculopathy develops over a longer horizon than was available for many members of the primary cohort, CAV analyses used a broader cohort with available longitudinal echocardiographic and angiographic surveillance data.

The broader CAV cohort did not require an eligible echocardiogram within the first post-transplant year. Only patients with available CAV grading could contribute to CAV analyses. Eligibility requirements differed according to whether the analysis evaluated first observed CAV or time-updated CAV status.

### 6. Longitudinal mixed-effects models

#### 6.1 Natural history of LV mass

The longitudinal course of LV mass was estimated using linear mixed-effects models. Fixed effects included linear time after transplantation and quadratic time after transplantation. The random-effects structure included a patient-specific random intercept and random slope for time, with correlation permitted between the random effects.

The recipient-sex model additionally included recipient sex as a fixed effect. Models were estimated using restricted maximum likelihood unless otherwise specified.

#### 6.2 Donor–recipient sex pairing

Donor–recipient sex pairing was categorized as:

- male donor–male recipient;
- female donor–female recipient;
- male donor–female recipient; and
- female donor–male recipient.

Because the four-category pairing variable incorporates recipient sex, recipient sex was not entered simultaneously in the sex-pairing model.

Interactions of sex pairing with linear and quadratic time were evaluated using likelihood-ratio testing. The main-effects and interaction models were fitted using maximum-likelihood estimation for this comparison. Because inclusion of the interaction terms did not improve model fit, final sex-pairing estimates were obtained from the main-effects model fitted using restricted maximum likelihood.

### 7. Bayesian joint models for graft dysfunction and mortality

#### 7.1 Model configuration

Separate Bayesian joint models were fitted for graft dysfunction and mortality. Each joint model included a patient-level time-to-event dataset and a corresponding repeated-measures LV mass dataset constructed using the event-specific temporal restrictions described above.

#### 7.2 Longitudinal submodel

The longitudinal submodel was a linear mixed-effects model of LV mass divided by 10. Fixed effects included:

- linear time after transplantation;
- quadratic time after transplantation; and
- recipient sex.

The random-effects structure included a patient-specific random intercept and random slope for time, with correlation permitted between the two random effects.

The quadratic fixed effect allowed the mean LV mass trajectory to be nonlinear. The random intercept represented persistent patient-level deviation from the expected LV mass level, whereas the random slope represented patient-level deviation from the expected rate of LV mass change.

The initial longitudinal model was fitted using the nlme package.

#### 7.3 Survival submodel

The survival submodel used time from transplantation to graft dysfunction, death, or censoring. Recipient sex was included as a baseline covariate.

#### 7.4 Longitudinal–survival association structure

The longitudinal and survival submodels were linked through two features of the underlying model-estimated LV mass trajectory:

- the current fitted LV mass at each time point; and
- the instantaneous rate of change in fitted LV mass at that time.

The current-value and slope association terms were included simultaneously. Each association was therefore estimated while accounting for the other component of the LV mass trajectory.

Current-value hazard ratios were reported per 10-g higher underlying LV mass. Slope hazard ratios were reported per 1-g/year.

#### 7.5 Bayesian estimation and convergence

Joint models were fitted using JMbayes2. Final models used:

- 60,000 Markov chain Monte Carlo iterations;
- 15,000 burn-in iterations;
- a thinning interval of 5;
- six parallel chains;
- four computational cores; and
- random-number seed 456.

Posterior means and 95% credible intervals were extracted for the current-value and slope association parameters and exponentiated to obtain hazard ratios.

Convergence was assessed using trace plots, posterior density plots, potential scale-reduction diagnostics, and parameter-specific R-hat values. Fitted model objects, summaries, coefficient structures, and diagnostic plots were retained for audit.

#### 7.6 Expanded and indexed sensitivity analyses

Expanded joint models additionally adjusted the survival submodel for:

- recipient age at transplantation;
- donor age; and
- recipient serum creatinine at transplantation.

The primary joint models were also repeated using LV mass indexed to recipient body surface area. In indexed analyses, current-value effects were reported per 10-g/m² higher LV mass index and slope effects per 1-g/m²/year greater LV mass index slope.

### 8. Remodeling phenotype

#### 8.1 Score construction

A patient-level LV mass remodeling score was constructed from the primary graft-dysfunction joint model. The score combined each patient’s estimated deviation from the expected LV mass level and estimated deviation from the expected LV mass slope with the corresponding joint-model association coefficients.

Specifically, the patient-specific random intercept was multiplied by the posterior current-value association coefficient, and the patient-specific random time slope was multiplied by the posterior slope association coefficient. The two weighted components were then added:

Remodeling score_i_ = α_value_b_0i_+α_slope_b_1i_,

where b_0i_ is the patient-specific random intercept, b_1i_ is the patient-specific random slope, and the α terms are the corresponding association coefficients from the graft-dysfunction joint model.

Patients were divided at the cohort median into adverse and non-adverse remodeling groups.

Because the remodeling phenotype was derived from the same joint model used to estimate the outcome association, group comparisons were intended to provide a clinically interpretable description of the modeled associations rather than independent prognostic validation.

#### 8.2 Description of group-specific trajectories

For descriptive interpretation, patient-specific estimated LV mass at transplantation and patient-specific linear LV mass slope were calculated from the fixed and random effects of the longitudinal mixed model.

Remodeling groups were compared using Kaplan–Meier curves, log-rank tests, and Cox proportional-hazards models for graft dysfunction and mortality. Inferential analyses used all available follow-up.

#### 8.3 Echocardiographic surveillance comparison

To evaluate whether remodeling classification could have been influenced by differences in echocardiographic surveillance, adverse and non-adverse remodeling groups were compared with respect to:

- total number of eligible echocardiograms;
- number of eligible echocardiograms during the first year;
- time from first to last eligible echocardiogram;
- annualized imaging frequency;
- timing of the first eligible echocardiogram; and
- timing of the last eligible echocardiogram.

### 9. Exploratory joint model for CAV

A separate Bayesian joint model evaluated the first observed CAV grade 2 or 3. Follow-up began when the patient first had eligible longitudinal LV mass observation. Patients without the event were censored at their last available angiogram.

LV mass measurements were required to occur before the CAV event or censoring time. For patients who developed CAV grade 2 or 3, LV mass measurements during the 90 days immediately preceding the event were excluded. At least two eligible LV mass measurements were required.

The longitudinal submodel included linear and quadratic time after transplantation, recipient sex, a patient-specific random intercept, and a patient-specific random slope. The CAV survival submodel included recipient sex. Current fitted LV mass and instantaneous LV mass slope were entered simultaneously in the association structure.

The CAV joint model used:

- 60,000 iterations;
- 15,000 burn-in iterations;
- thinning of 5;
- six chains;
- four computational cores; and
- random-number seed 789.

The CAV joint model was considered exploratory because angiographic surveillance was less frequent than echocardiographic surveillance and occurred at clinically variable times.

### 10. Time-updated Cox models

#### 10.1 Graft dysfunction and subsequent mortality

Graft dysfunction was evaluated as a time-varying exposure in a counting-process Cox proportional-hazards model.

Patients without graft dysfunction before death or censoring contributed one interval classified as free of graft dysfunction. Patients who developed graft dysfunction before death or censoring contributed:

- a pre-graft-dysfunction interval; and
- a post-graft-dysfunction interval beginning on the graft-dysfunction date.

The primary model evaluated mortality as a function of time-updated graft-dysfunction status and recipient sex. Robust standard errors were clustered by patient to account for multiple intervals contributed by the same individual.

A subsequent model additionally included the continuous remodeling score to evaluate whether graft dysfunction remained associated with mortality after accounting for the LV mass remodeling phenotype.

#### 10.2 CAV and subsequent mortality

CAV grade 2 or 3 was evaluated as a time-varying exposure in a counting-process Cox model using the broader angiographic-surveillance cohort.

Follow-up began at the first available angiogram. Patients were classified as free of CAV grade 2 or 3 until the date of the first qualifying angiogram and as having CAV thereafter.

Patients who developed CAV contributed separate pre-CAV and post-CAV follow-up intervals. Patients who did not develop CAV contributed a single interval. The model included time-updated CAV status and recipient sex, with robust standard errors clustered by patient.

### 11. Factors associated with longitudinal LV mass

#### 11.1 General modeling strategy

Candidate factors associated with LV mass were evaluated using linear mixed-effects models estimated by restricted maximum likelihood.

All models included:

- linear time after transplantation;
- quadratic time after transplantation;
- recipient sex;
- a patient-specific random intercept; and
- a patient-specific random slope for time.

When the random-intercept and random-slope structure could not be estimated reliably, a random-intercept-only model was used and the final random-effects structure was recorded.

Each candidate exposure was first evaluated in a separate model using the common adjustment structure. These analyses were intended to characterize exposure-specific associations with LV mass and were not interpreted as mutually independent or causal effects.

Effect estimates were reported as adjusted differences in LV mass in grams with 95% confidence intervals and two-sided p values. LV mass was divided by 10 during model estimation when required for numerical scaling, and coefficients and confidence intervals were multiplied by 10 for presentation in grams.

#### 11.2 Time-invariant characteristics

Time-invariant characteristics included donor, recipient, and transplant variables measured at or before transplantation, including:

- donor and recipient age;
- donor height and weight;
- recipient body mass index;
- donor–recipient sex pairing;
- primary cardiac diagnosis;
- race and ethnicity;
- blood group;
- waitlist and transplant characteristics;
- donor medical history;
- donor terminal laboratory values;
- donor mechanism and circumstance of death; and
- donor and recipient viral serostatus.

Each characteristic was entered separately into the common mixed-effects model.

For continuous variables, coefficients represent the average difference in LV mass associated with the specified unit increase. For categorical variables, coefficients represent the average difference from the designated reference category over follow-up.

Categories containing fewer than 10 patients were combined into an “other” category when clinically appropriate. When no reference category was prespecified, the most frequently represented category was generally selected.

#### 11.3 Time-varying exposures

Time-varying exposures included:

- blood pressure measurements;
- routine laboratory values;
- cardiac biomarkers;
- immunosuppressant trough concentrations;
- donor-specific antibodies;
- calculated panel-reactive antibody;
- conventional biopsy rejection measures; and
- Molecular Microscope Diagnostic System transcript scores.

For each echocardiographic LV mass measurement, the closest eligible preceding exposure measurement within the prespecified lookback window was selected.

The lookback windows were:

- 90 days for routine laboratory measurements, cardiac biomarkers, and immunosuppressant trough concentrations;
- 180 days for vital signs, conventional biopsy findings, and MMDx measurements; and
- 365 days for donor-specific antibodies and calculated panel-reactive antibody.

Laboratory, blood-pressure, and immunosuppressant measurements could occur on the same day as or before the echocardiogram. Conventional biopsy and MMDx measurements were required to precede the echocardiogram.

#### 11.4 Between-patient and within-patient decomposition

Each time-varying exposure was decomposed into a between-patient and within-patient component.

For patient (i), the between-patient component was the patient-specific mean exposure: $\bar{x_{i}}$

The within-patient component for observation (j) was: $x_{ij}-\bar{x_{i}}$

Both terms were entered simultaneously in the mixed-effects model.

The between-patient coefficient estimates whether patients with higher average exposure had higher or lower average LV mass than patients with lower average exposure. The within-patient coefficient estimates whether LV mass was higher or lower when a patient’s current exposure differed from that patient’s own usual exposure level.

Patients with no variation in an exposure could contribute to estimation of the between-patient association but did not directly contribute information to the within-patient association. A within-patient estimate was reported only when at least five patients demonstrated within-patient variation in the exposure.

#### 11.5 Blood-pressure measurements

Systolic blood pressure, mean arterial pressure, and pulse pressure were evaluated separately.

Pulse pressure was calculated as:

Pulse pressure=Systolic blood pressure−Diastolic blood pressure.

Mean arterial pressure was calculated as:

Mean arterial pressure=Diastolic blood pressure+1/3(Pulse pressure).

Blood-pressure measurements were modeled per 10-mmHg increase using the closest measurement obtained on or before the echocardiogram and within the preceding 180 days.

#### 11.6 Laboratory measurements and cardiac biomarkers

Routine laboratory and biomarker values were matched using the closest measurement obtained on or before the echocardiogram and within the preceding 90 days.

Values were scaled in clinically interpretable units. Blood urea nitrogen, glucose, alkaline phosphatase, aspartate aminotransferase, and alanine aminotransferase were modeled per 10-unit increase. Total cholesterol, high-density lipoprotein cholesterol, and low-density lipoprotein cholesterol were modeled per 10-mg/dL increase.

NT-proBNP, high-sensitivity troponin T, creatine kinase, and triglycerides were transformed using:

log_2_(x+1)

The corresponding coefficient was interpreted as the approximate difference in LV mass associated with a doubling of the biomarker plus one.

Each laboratory exposure was evaluated in a separate between-patient and within-patient model.

#### 11.7 Immunosuppressant trough concentrations

Tacrolimus, sirolimus, and available everolimus trough concentrations were evaluated separately. Concentrations were modeled per 1-ng/mL increase using the closest result obtained on or before the echocardiogram and within the preceding 90 days.

The primary analyses used between-patient and within-patient decomposition.

#### 11.8 Donor-specific antibodies and calculated panel-reactive antibody

Donor-specific antibodies and calculated panel-reactive antibody were evaluated separately for HLA class I and class II.

Baseline exposure was defined using the closest available measurement obtained on or before transplantation. For longitudinal analyses, the closest post-transplant measurement obtained on or before each echocardiogram and within the preceding 365 days was selected.

When multiple measurements occurred on the same day, cPRA was represented by the maximum percentage and DSA was classified as positive when any result was positive.

cPRA was modeled per 10-percentage-point increase. DSA was modeled as positive versus negative. Longitudinal measurements were evaluated using between-patient and within-patient decomposition.

#### 11.9 Conventional rejection

Conventional rejection exposures were defined using the closest preceding endomyocardial biopsy within 180 days of each LV mass measurement. Same-day biopsies were excluded.

Acute cellular rejection was defined as International Society for Heart and Lung Transplantation grade 1R or higher. Antibody-mediated rejection activity was represented by the percentage of positive immunostaining and was modeled per 10-percentage-point increase.

When no eligible biopsy had been performed during the preceding 180 days, both rejection measures were coded as zero, representing no observed rejection exposure during the lookback period. When a biopsy had been performed but the rejection grade or immunostain result was unavailable, the corresponding exposure remained missing. Because absence of a biopsy was coded as absence of rejection, rejection exposures reflect documented rejection during the lookback period rather than verified rejection-free status, and the resulting estimates should be interpreted accordingly.

Each rejection measure was decomposed into a patient-specific mean and a within-patient deviation. For binary ACR, the patient-specific mean represented the proportion of eligible observations associated with ACR grade 1R or higher. For immunostaining, the patient-specific mean represented the average immunostain burden.

#### 11.10 Molecular microscope measurements

For each LV mass measurement, the closest MMDx assessment occurring within the preceding 180 days was selected. Same-day MMDx assessments were excluded.

Continuous MMDx scores were standardized to a mean of zero and standard deviation of one. Effect estimates therefore represent the adjusted difference in LV mass associated with a 1-standard-deviation higher score.

Each continuous MMDx score was evaluated in a separate between-patient and within-patient model. These models additionally adjusted for the interval between the MMDx assessment and echocardiogram, expressed in 30-day units.

Categorical MMDx interpretations were classified as normal, slightly abnormal, or abnormal, with normal used as the reference. Separate indicators were created for slightly abnormal and abnormal interpretations and decomposed into between-patient and within-patient components.

#### 11.11 Multivariable determinants model

A multivariable mixed-effects model was constructed using variables selected a priori on clinical grounds and representing nonredundant recipient, donor, hemodynamic, rejection, renal, and metabolic domains.

The final model simultaneously included:

- recipient sex;
- recipient age at transplantation;
- recipient body mass index at transplantation;
- donor age;
- donor height;
- donor weight;
- donor terminal creatinine;
- between-patient and within-patient pulse pressure;
- between-patient and within-patient ACR;
- between-patient and within-patient immunostain burden;
- between-patient and within-patient recipient creatinine; and
- between-patient and within-patient anion gap.

The model included linear and quadratic time after transplantation and patient-specific random intercepts and slopes. No automatic variable-selection procedure was used.

### 12. Descriptive LV mass trajectory analyses

The overall post-transplant LV mass trajectory was summarized in approximately monthly time bins. Within each bin, the number of measurements, number of contributing patients, mean, median, and interquartile range of LV mass were calculated. Bins represented by fewer than 10 patients were excluded.

### 13. Transcriptomic validation cohort

**13.1 Sample collection and storage**

Biopsy samples were collected during protocol surveillance biopsies under a consented research protocol. Samples were placed in RNAlater and transferred immediately to a −80°C freezer at Houston Methodist Hospital, then shipped under maintained cold storage to Northwestern University under a material transfer agreement.

**13.2 RNA extraction and library preparation**

Total RNA was extracted from frozen biopsies using the Quick-RNA Microprep Kit (Zymo Research) with ZR BashingBead Lysis tubes (Zymo Research). RNA was further concentrated and purified using the RNA Clean and Concentrator kit (Zymo Research). cDNA libraries were generated using the NEBNext Single Cell/Low Input cDNA Synthesis and Amplification Module (New England Biolabs).

**13.3 Sequencing**

Approximately 200 fmol of cDNA from each sample was barcoded and pooled using the Native Barcoding Kit 24 V14 (Oxford Nanopore Technologies), with 7-12 samples per pool. Approximately 20 fmol of each pool was loaded onto an R10.4.1 PromethION flow cell (Oxford Nanopore Technologies) and sequenced on an Oxford Nanopore PromethION platform. Data acquisition and base calling were performed using MinKNOW.

**13.4 Quantification and differential expression**

Raw sequencing reads were aligned to the GRCh38 human reference genome, and gene expression was quantified using featureCounts. TMM normalization factors were calculated using edgeR, and the mean-variance relationship was modeled with voom to generate observation-level precision weights. Associations between gene expression and the patient-level remodeling score, modeled as a continuous variable, were tested using limma-voom with empirical Bayes moderation. The model adjusted for biopsy timing and donor sex. Because some patients contributed more than one biopsy, repeated samples from the same patient were accounted for using duplicate correlation, with patient identifier as the blocking variable.

**13.5 Gene set enrichment analysis**

Gene set enrichment analysis was performed using all tested genes ranked by the moderated t statistic against the MSigDB Hallmark and Gene Ontology Biological Process, Cellular Component, and Molecular Function collections. Positive normalized enrichment scores indicated enrichment with increasing remodeling score, whereas negative scores indicated enrichment with decreasing remodeling score. A Benjamini-Hochberg false discovery rate below 0.05 was considered statistically significant. Full enrichment results are reported in Supplementary Table S5.

### 14. Missing data and quality control

No general statistical imputation was performed. Each analysis used observations with complete data for the variables required by that model.

Quality-control procedures included the following:

- event and censoring times were required to be positive;
- LV mass measurement times could not exceed the corresponding event or censoring time;
- patients with graft dysfunction on or before post-transplant day 90 and all their echocardiograms were excluded from the primary remodeling cohort;
- patients with an event had to satisfy the prespecified 91-day LV mass-to-event lead time;
- at least two eligible LV mass observations were required for joint-model analyses;
- identifiers in longitudinal and survival datasets were realigned after complete-case filtering;
- patient counts, event counts, measurements per patient, measurement timing, and survival times were summarized for each analytic cohort;
- duplicate patient identifiers and conflicting patient-level characteristics were examined;
- model-specific datasets, audit tables, fitted model objects, diagnostic plots, and software-session information were retained.

### 15. Statistical software and reporting

Data preparation was performed in Python using pandas and numpy. Statistical analyses were performed in R version 4.5.2 using nlme, survival, JMbayes2, survminer, and ggplot2, as applicable.

Linear mixed-effects models were estimated using restricted maximum likelihood except when maximum-likelihood estimation was required for nested model comparisons.

Frequentist Cox models reported hazard ratios with 95% confidence intervals and two-sided p values. Robust variance estimates were used for counting-process models in which patients contributed multiple follow-up intervals.

Bayesian joint models reported posterior hazard ratios with 95% credible intervals. Current-value associations were presented per 10-g higher underlying LV mass, and slope associations were presented per 1-g/year greater LV mass slope after rescaling.

Kaplan–Meier comparisons used log-rank tests, with Cox models used for effect estimation. Remodeling-group comparisons, descriptive trajectory analyses, pre-event summaries, and CAV analyses were interpreted as descriptive or exploratory where specified. Analyses of candidate factors associated with LV mass involved a large number of exposures and were not adjusted for multiple comparisons; these estimates are hypothesis-generating. Two-sided p values below 0.05 were considered statistically significant.
